# Delivering HIV pre-exposure prophylaxis (PrEP) care online: A scoping review

**DOI:** 10.1101/2021.12.15.21267774

**Authors:** Ross Kincaid, Claudia Estcourt, Jamie Frankis, Jenny Dalrymple, Jo Gibbs

## Abstract

**Objectives:** HIV pre-exposure prophylaxis (PrEP), in which people take HIV medication to prevent HIV acquisition, is a highly effective method of HIV prevention; however, global implementation of PrEP is patchy. PrEP provision will need to be upscaled significantly to achieve UNAIDS/WHO goals of elimination of HIV transmission. Online provision of PrEP care could enhance access to, and delivery of, care at scale. We explored the extent to which PrEP care has been delivered online to inform the development of a novel online PrEP clinic.

**Design:** Scoping review.

**Data sources:** Embase, MEDLINE, Web of Science, CINAHL, PsycINFO, ASSIA, PUBMED, Open Grey, and EThOS databases.

**Eligibility criteria:** English language articles describing a service that delivered one or more element of PrEP-related care online, published from 2009 onwards.

**Data extraction and synthesis:** Data were extracted using matrices and synthesised using summary statistics and thematic analysis. The Mixed Methods Appraisal Tool was used to assess study quality.

**Results:** Fifty-nine articles were included: eight randomised controlled trials, 12 non-randomised quantitative studies, 30 descriptive quantitative studies, 14 qualitative studies, and four reviews of online content. Seven studies detailed comprehensive PrEP care pathways that used a combination of online, face-to-face and telephone based care. Of the remaining studies, the majority focused on HIV testing outside a PrEP context. Care tended to be delivered via websites (n=41), video chat, and smartphone apps (both n=10). The acceptability and feasibility of delivering elements of care online was high.

**Conclusions:** Online PrEP care appears feasible and acceptable, offering convenience and a means to overcome some of the reported barriers to face-to-face care. Services tended to focus on a single element of PrEP-related care or use a combination of online, face-to-face and phone-based care. Additional formative work is needed to inform the development of complete online PrEP care pathways.

**Strengths and limitations of this study:**

- This scoping review provides a comprehensive, critical overview of existing literature related to online provision of HIV pre-exposure prophylaxis (PrEP)-related care.
- We developed a comprehensive definition of PrEP care which took into consideration all essential elements of care required for safe and appropriate PrEP prescribing.
- Established guidelines for conducting and reporting scoping reviews were followed throughout.
- We conducted an exhaustive search of nine databases.
- The heterogeneity in design and aim of the included studies limited our ability to synthesise findings.

## INTRODUCTION

HIV remains an important health concern.[1] Oral HIV pre-exposure prophylaxis (PrEP), in which people take HIV medication to prevent HIV acquisition, has been found to reduce HIV transmission by up to 86%.[2-4] However, global implementation of PrEP is patchy and has proved challenging in some settings.[5, 6] The UNAIDS 2020 HIV prevention road map states that a lack of systematic implementation of HIV prevention methods at scale is slowing efforts to reach targets.[7-9]. Only one third of the target 3 million people had started PrEP by the end of 2020.[7, 9]

PrEP provision (PrEP care) involves several elements of care: baseline testing for HIV and other blood borne viruses, sexually transmitted infection (STI) screening as appropriate, renal function assessment, and prescribing of a fixed formulation of antiviral drugs, most commonly tenofovir disoproxil and emtricitabine.[10, 11] Regular monitoring is recommended in many countries’ guidelines and includes adherence support, repeat infection screening, and renal function monitoring.[10, 11] PrEP care is provided in a variety of settings including sexual health services, HIV services, and community settings. Interventions which provide PrEP care more efficiently could play an important role in scaling up PrEP coverage and, ultimately, meeting UNAIDS HIV elimination targets.[12] Digital health interventions, which are increasingly incorporated into sexual healthcare could offer such a solution.[13, 14]

We sought to understand the extent to which elements of PrEP-related care had been delivered online to inform the development of a novel online PrEP service (the ePrEP clinic). The ePrEP clinic aims to deliver a complete PrEP pathway online (i.e. online consultation, remote sampling, and electronic PrEP prescription). We undertook a scoping review rather than a systematic review because of the novelty of the online approach to PrEP provision and the heterogeneity of the existing literature. Specifically, we aimed to establish what, and how, elements of PrEP care have been delivered online, the feasibility and acceptability of these services, and what barriers to, and facilitators of, online PrEP care have been identified.

## METHODS

We used the Arksey *et al*. and Levac *et al*. frameworks for conducting scoping reviews and used the PRISMA extension for Scoping Reviews to report the findings.[15-17] The protocol for this review can be accessed here: https://doi.org/10.6084/m9.figshare.17157899.

### Defining PrEP care and search terms

We developed a comprehensive definition of PrEP care in order to identify search terms for the review. The joint BHIVA BASHH recommendations for PrEP use were used as a basis for this definition.[10] We chose to focus on elements of PrEP care that would be essential for an online PrEP service to provide safe and appropriate care (herein referred to as PrEP-related elements of care): HIV testing, renal function monitoring, PrEP prescription, PrEP eligibility/HIV risk assessment, PrEP education, PrEP adherence monitoring and support, and monitoring of PrEP-related side-effects and possible drug interactions. A full overview of the process for developing our definition of PrEP care is found in **Supplementary Material 1**.

We established search terms by, where possible, adapting terms used in existing systematic reviews that focused on HIV testing, renal function monitoring, and digital health [18-20] The PrEP-related elements of care were split into four groups given the complexity of the search; these groups formed four independent searches: HIV testing; renal function monitoring; PrEP prescription; and general PrEP care. The full terms can be found in **Supplementary Material 1**.

### Search strategy

Initial searches were conducted in February 2019. An updated search was conducted on the 28^th^ of July 2021 with the same search strategy. EMBASE, MEDLINE, Web of Science, CINAHL, ASSIA, PsycINFO, PubMed, Open Grey, and British Library EThOS were searched using the terms outlined in **Supplementary Material 1**. Open Grey was no longer accessible at the time of the updated search as it ceased to be an active resource. Additional records were sought by searching the reference list of eligible articles – no additional records were identified. The PRISMA flow diagram can be found in **Figure 1**. RK conducted the searches and screening; any uncertainties were checked by JG.

**Figure 1.**
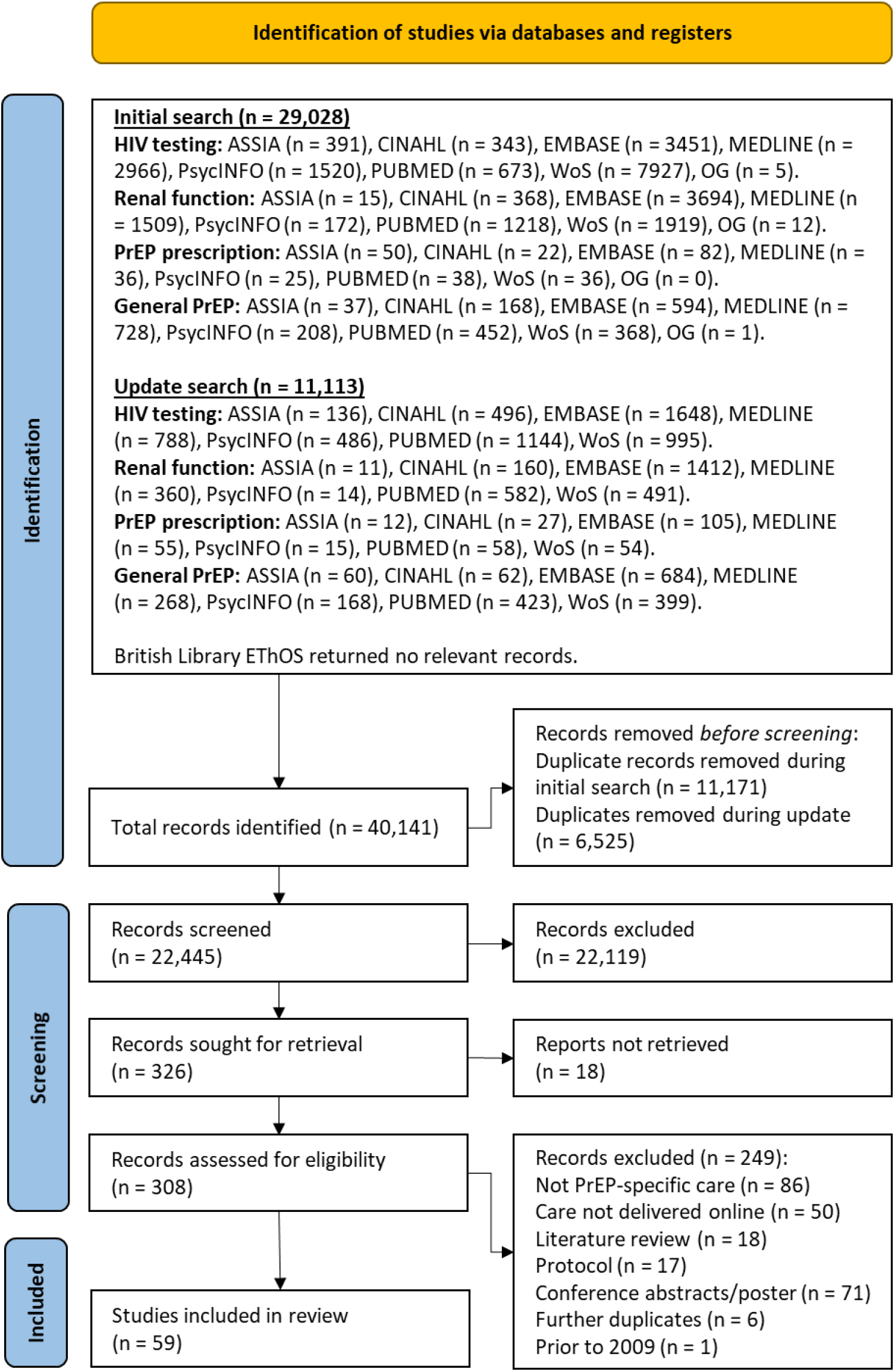
PRISMA flow diagram for the initial and updates searches.

### Eligibility criteria

Eligible articles were published or made available between 01.01.2009 and 28.07.2021 and written in English. Articles had to describe a service that delivered at least one PrEP-related element of care online (though this did not have to be within the context of PrEP care; e.g. ordering an HIV self-test kit online). The study had to target patients or clients directly. Studies where the internet was required to download an app but where that app was used entirely offline were excluded as were studies that examined training or support to healthcare professionals online. Studies were excluded if they focused solely on SMS or only recruited potential patients/clients without providing actual care. Conference abstracts, posters, presentations, and literature reviews were excluded from this review. Reviews of online content (e.g. websites and YouTube videos) were included if the content reviewed met the other eligibility criteria. The criteria are summarised in Box 1.

#### Box 1: Summary of inclusion and exclusion criteria.

##### Summary of inclusion and exclusion criteria

###### Inclusion criteria

- Studies which delivered one or more element of PrEP-related care online.
- Studies that targeted patients or service users directly.
- Studies written in English and published or made available between 01.01.2009 and 28.07.2021.

###### Exclusion criteria

- Studies where the internet was required to download an app but where that app was used entirely offline.
- Studies that focused solely on SMS.
- Studies that only recruited or identified potential patients but which did not actually deliver care.
- Studies that focused on healthcare professionals (e.g. online support or training).
- Conference abstracts, protocols, posters, and literature reviews.

### Selection of sources of evidence

Articles’ titles and abstracts were screened in Mendeley.[21] Full texts were sourced for eligible studies and data were extracted into a data extraction table created in Microsoft Excel (see **Supplementary Material 2** for a template of this table).[22]

### Analysis of included studies

The Mixed Methods Appraisal Tool was used to assess the quality of the included articles given the variation in study design.[23] Matrices were then used to map out the extracted data and, where appropriate, thematic analysis was used to identify key themes across the included studies.

### Patient and public involvement

No patients or members of the public were involved in conducting this scoping review.

## RESULTS

The initial searches identified 29,028 articles and the updated searches identified a further 11,113. Three hundred and eight full text articles were assessed of which 59 met the inclusion criteria. The included studies had sample sizes ranging from 11 to 19,497 for quantitative studies, and 10 and 59 for qualitative studies.[24-27] The articles were published between 2012 and 2021 and the number published per year seem to follow an upward trend until 2020 – possibly disrupted by the COVID-19 pandemic. Studies were predominantly conducted in the USA (n=27, 45.8%), nine (15.3%) were conducted in the UK, seven (11.9%) in Canada, six (10.2%) in China, three (5.1%) in Thailand and one (1.7%) in each of Brazil, France, Italy, Netherlands, and South Korea.

Study design was not consistently reported and so we adopted the categories used in the MMAT to group study designs to aid clear and complete reporting.[23] Eight articles (13.6%) outlined randomised controlled trials, 12 (20.3%) were categorised as non-randomised quantitative studies, 30 (50.8%) were categorised as descriptive quantitative studies, and 14 (23.7%) used qualitative methods. Of the 14 qualitative studies, eight implemented focus groups and eight used semi-structured interviewing – two studies used both methods. Four articles reviewed existing online content. A total of nine studies used a mixed-methods design which accounts for the total designs exceeding the number of studies. The quality of the included studies varied – see **Supplementary Material 3** for the full MMAT assessment. It was not possible to meaningfully synthesise studies’ outcomes due to the heterogeneity of the studies’ designs and objectives. Instead, the main findings of the included studies have been reported in **Supplementary Material 4**. We do, however, provide condensed overviews of the included articles below and comment on the feasibility and acceptability of included studies below which was the aim of some of these studies.

### Which elements of PrEP care have been delivered online, and how?

PrEP-related elements of care were mainly delivered via websites (n=41), followed by video call (n=10), smartphone apps (n=10), email (n=6), and YouTube videos (n=2), with some studies implementing more than one modality. Sixteen studies delivered PrEP-related elements of care in the context of PrEP,[24, 28-42] 38 delivered an aspect of HIV testing online without being explicitly related to PrEP,[25-27, 43-77] one study provided renal test results online outside a PrEP context,[78] and four studies reviewed existing online content.[79-82] We have grouped our findings depending on whether or not PrEP is explicitly incorporated into each service given the possible higher relevance of the studies that delivered care in a PrEP context.

#### Services within a PrEP context

There was considerable heterogeneity in the type and number of PrEP-specific care components delivered in the 16 studies that delivered PrEP-related care in the context of PrEP. Aspects of HIV testing were delivered online in ten studies, specifically: ordering HIV self-tests (HIVSTs) or self-sampling kits (n=4), online booking for a face-to-face appointment (n=2), providers sending results to patients (n=2), patients sending results to providers (n=1), online HIV counselling (n=1), and an online instructional video for completing HIVSTs (n=1). Two studies provided services relating to renal monitoring; specifically, one allowed participants to order a self-sampling kit for creatinine analysis,[29] and the other offered online booking for in-person renal function tests.[34] One service, documented in two papers, allowed participants to order 90 days’ worth of PrEP online following satisfactory test results and assessment.[35, 36] An online PrEP eligibility assessment or HIV risk assessment was offered in eight studies, PrEP education was provided online in nine studies, and eight studies either monitored or provided support for PrEP adherence online. No studies explicitly mentioned that they allowed participants to monitor PrEP side-effects and/or possible drug interactions online. Table 1 provides an overview of the studies that explicitly involved PrEP.

**Table 1.**
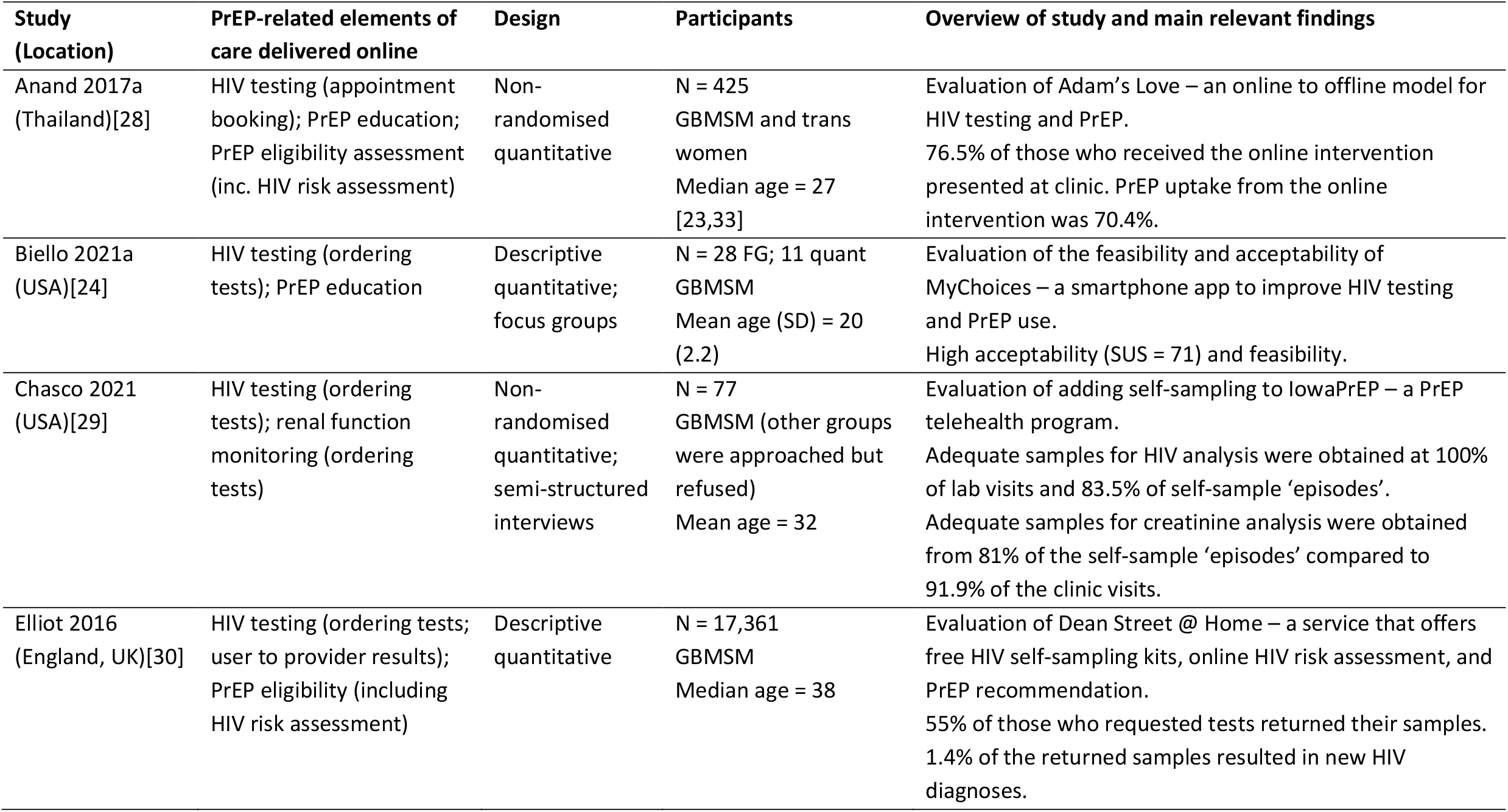
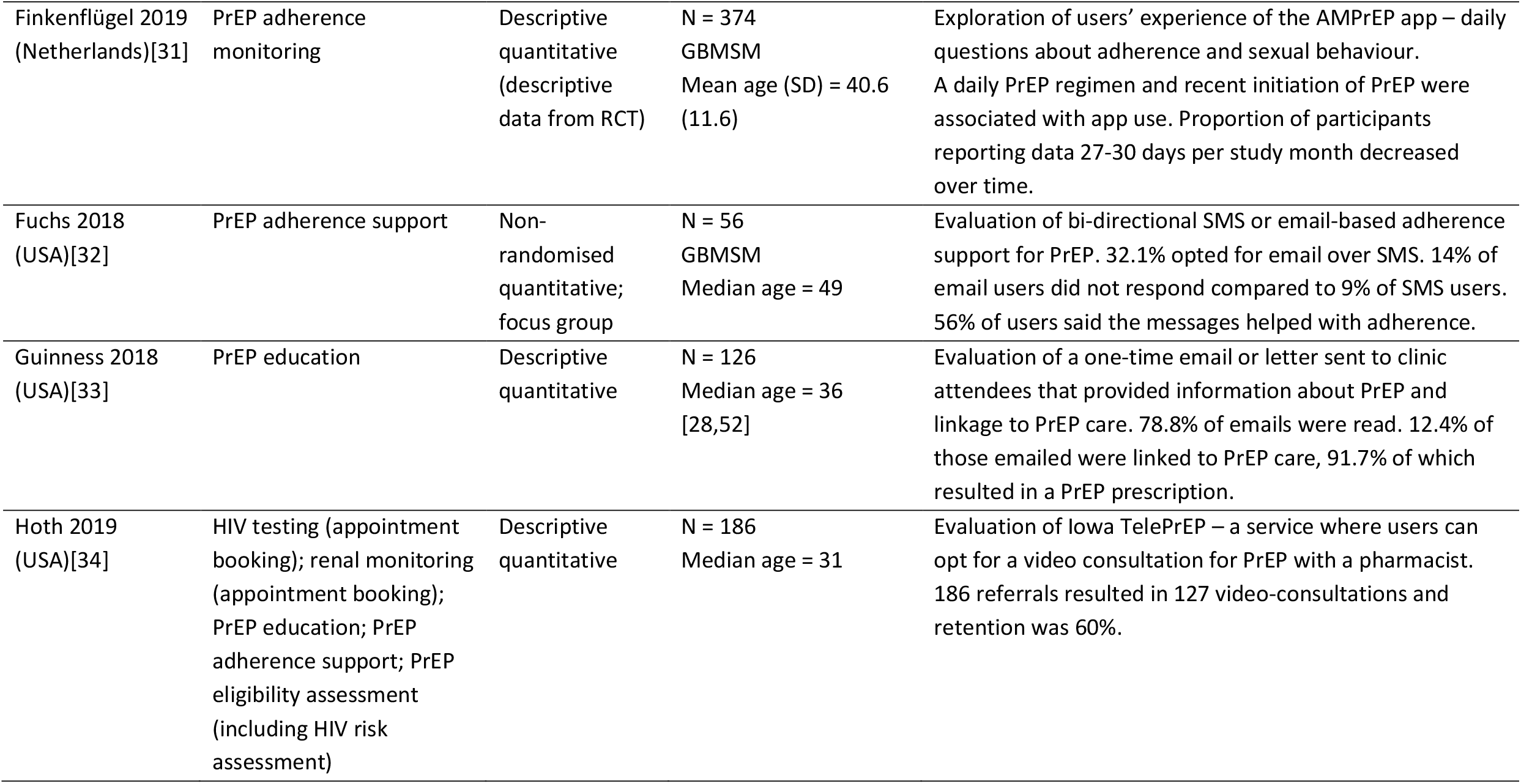
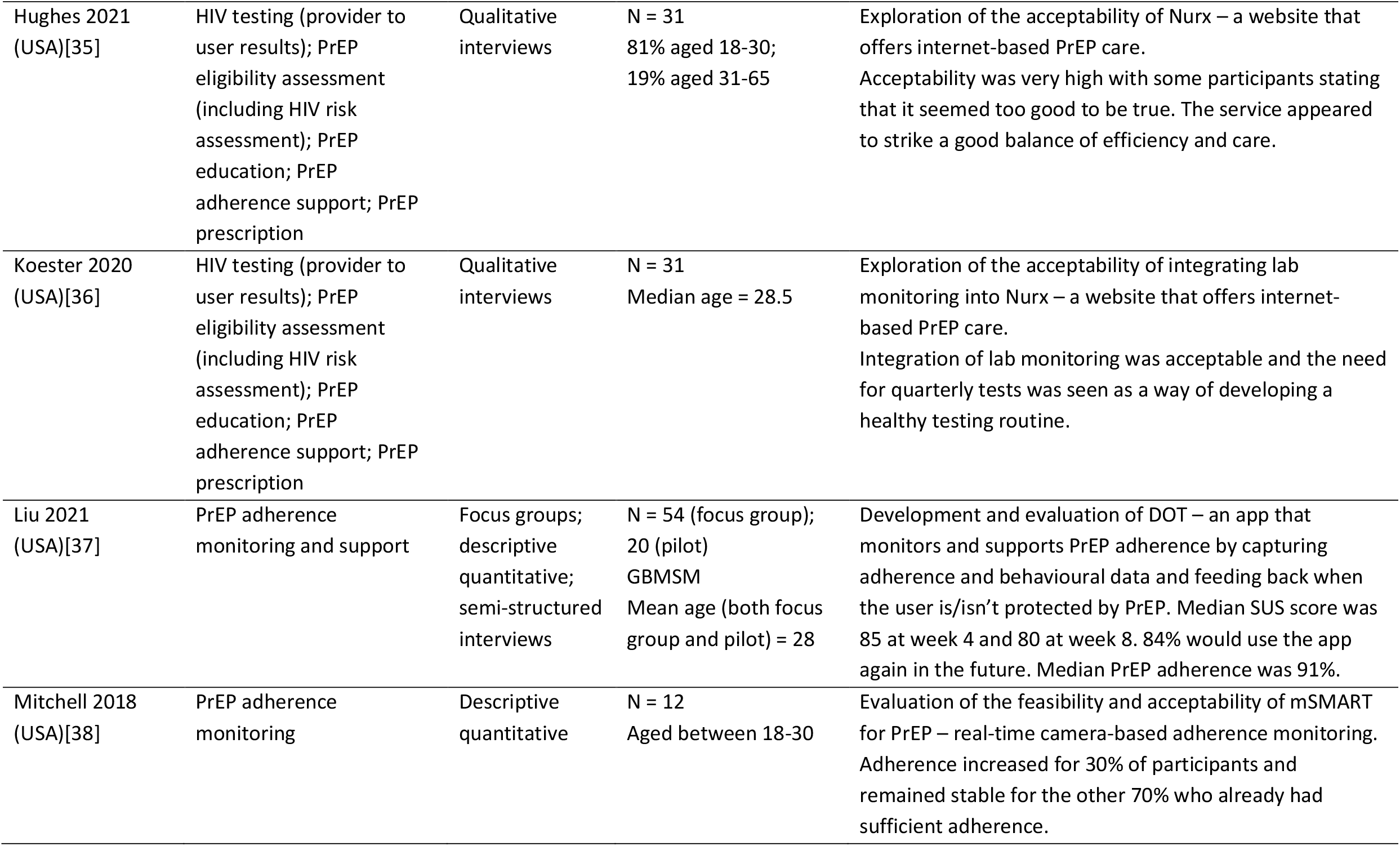
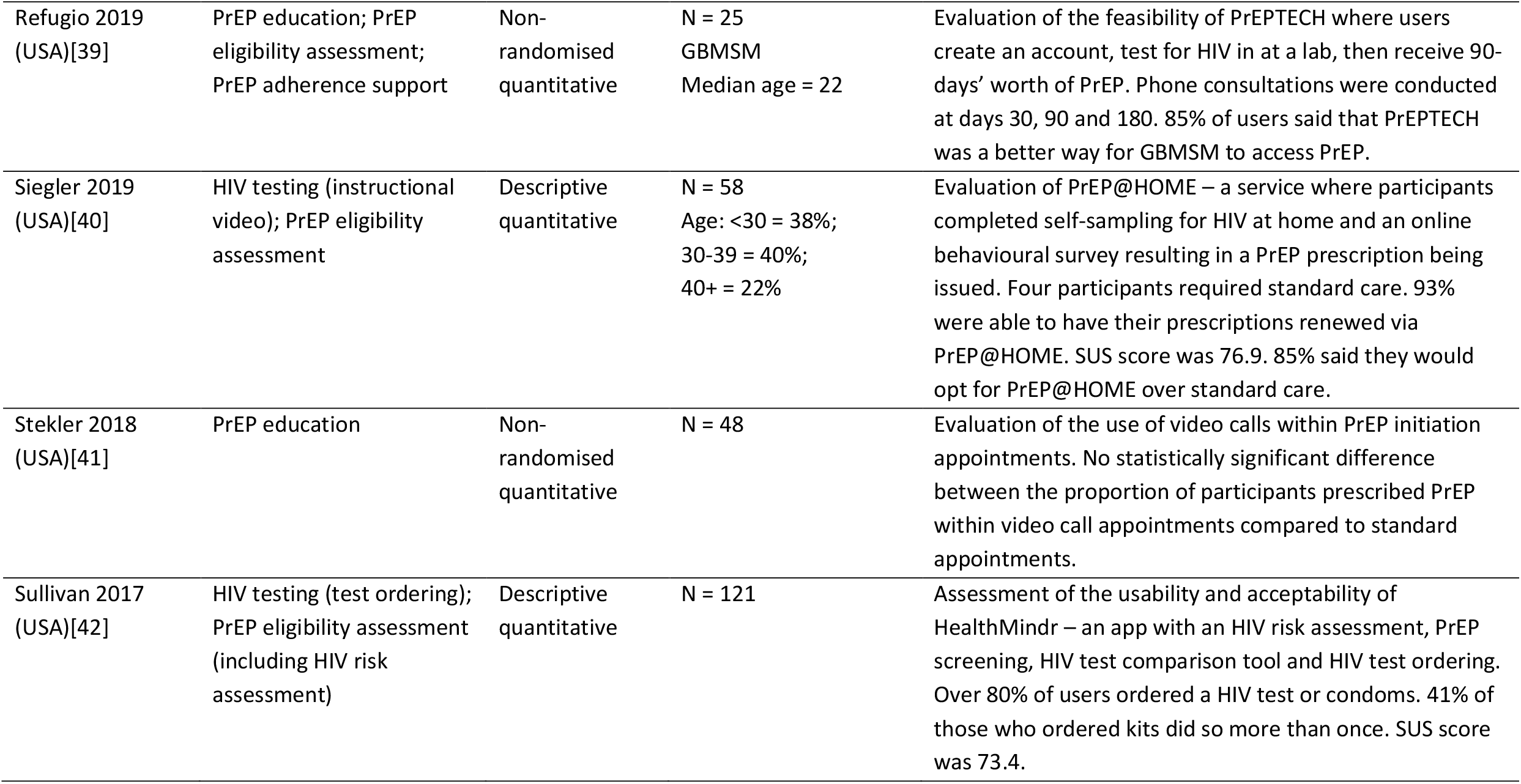
Overview of studies that explicitly involved PrEP.

| Study<br>(Location) | PrEP-related elements of<br>care delivered online | Design | Participants | Overview of study and main relevant findings |
| --- | --- | --- | --- | --- |
| Anand 2017a<br>(Thailand)[28] | HIV testing (appointment<br>booking); PrEP education;<br>PrEP eligibility assessment<br>(inc. HIV risk assessment) | Non-randomised<br>quantitative | N = 425<br>GBMSM and trans<br>women<br>Median age = 27<br>[23,33] | Evaluation of Adam's Love – an online to offline model for<br>HIV testing and PrEP.<br>76.5% of those who received the online intervention<br>presented at clinic. PrEP uptake from the online<br>intervention was 70.4%. |
| Biello 2021a<br>(USA)[24] | HIV testing (ordering<br>tests); PrEP education | Descriptive<br>quantitative;<br>focus groups | N = 28 FG; 11 quant<br>GBMSM<br>Mean age (SD) = 20<br>(2.2) | Evaluation of the feasibility and acceptability of<br>MyChoices – a smartphone app to improve HIV testing<br>and PrEP use.<br>High acceptability (SUS = 71) and feasibility. |
| Chasco 2021<br>(USA)[29] | HIV testing (ordering<br>tests); renal function<br>monitoring (ordering<br>tests) | Non-randomised<br>quantitative;<br>semi-structured<br>interviews | N = 77<br>GBMSM (other groups<br>were approached but<br>refused)<br>Mean age = 32 | Evaluation of adding self-sampling to IowaPrEP – a PrEP<br>telehealth program.<br>Adequate samples for HIV analysis were obtained at 100%<br>of lab visits and 83.5% of self-sample 'episodes'.<br>Adequate samples for creatinine analysis were obtained<br>from 81% of the self-sample 'episodes' compared to<br>91.9% of the clinic visits. |
| Elliot 2016<br>(England, UK)[30] | HIV testing (ordering tests;<br>user to provider results);<br>PrEP eligibility (including<br>HIV risk assessment) | Descriptive<br>quantitative | N = 17,361<br>GBMSM<br>Median age = 38 | Evaluation of Dean Street @ Home – a service that offers<br>free HIV self-sampling kits, online HIV risk assessment, and<br>PrEP recommendation.<br>55% of those who requested tests returned their samples.<br>1.4% of the returned samples resulted in new HIV<br>diagnoses. |

| Study (Location) | PrEP-related elements of care delivered online | Design | Participants | Overview of study and main relevant findings |
| --- | --- | --- | --- | --- |
| Finkenflügel 2019 (Netherlands)[31] | PrEP adherence monitoring | Descriptive quantitative (descriptive data from RCT) | N = 374<br>GBMSM<br>Mean age (SD) = 40.6 (11.6) | Exploration of users' experience of the AMPrEP app – daily questions about adherence and sexual behaviour. A daily PrEP regimen and recent initiation of PrEP were associated with app use. Proportion of participants reporting data 27-30 days per study month decreased over time. |
| Fuchs 2018 (USA)[32] | PrEP adherence support | Non-randomised quantitative; focus group | N = 56<br>GBMSM<br>Median age = 49 | Evaluation of bi-directional SMS or email-based adherence support for PrEP. 32.1% opted for email over SMS. 14% of email users did not respond compared to 9% of SMS users. 56% of users said the messages helped with adherence. |
| Guinness 2018 (USA)[33] | PrEP education | Descriptive quantitative | N = 126<br>Median age = 36 [28,52] | Evaluation of a one-time email or letter sent to clinic attendees that provided information about PrEP and linkage to PrEP care. 78.8% of emails were read. 12.4% of those emailed were linked to PrEP care, 91.7% of which resulted in a PrEP prescription. |
| Hoth 2019 (USA)[34] | HIV testing (appointment booking); renal monitoring (appointment booking); PrEP education; PrEP adherence support; PrEP eligibility assessment (including HIV risk assessment) | Descriptive quantitative | N = 186<br>Median age = 31 | Evaluation of Iowa TelePrEP – a service where users can opt for a video consultation for PrEP with a pharmacist. 186 referrals resulted in 127 video-consultations and retention was 60%. |
| Hughes 2021<br>(USA)[35] | HIV testing (provider to user results); PrEP eligibility assessment (including HIV risk assessment); PrEP education; PrEP adherence support; PrEP prescription | Qualitative interviews | N = 31<br>81% aged 18-30;<br>19% aged 31-65 | Exploration of the acceptability of Nurx – a website that offers internet-based PrEP care.<br>Acceptability was very high with some participants stating that it seemed too good to be true. The service appeared to strike a good balance of efficiency and care. |
| Koester 2020<br>(USA)[36] | HIV testing (provider to user results); PrEP eligibility assessment (including HIV risk assessment); PrEP education; PrEP adherence support; PrEP prescription | Qualitative interviews | N = 31<br>Median age = 28.5 | Exploration of the acceptability of integrating lab monitoring into Nurx – a website that offers internet-based PrEP care.<br>Integration of lab monitoring was acceptable and the need for quarterly tests was seen as a way of developing a healthy testing routine. |
| Liu 2021<br>(USA)[37] | PrEP adherence monitoring and support | Focus groups; descriptive quantitative; semi-structured interviews | N = 54 (focus group); 20 (pilot)<br>GBMSM<br>Mean age (both focus group and pilot) = 28 | Development and evaluation of DOT – an app that monitors and supports PrEP adherence by capturing adherence and behavioural data and feeding back when the user is/isn't protected by PrEP. Median SUS score was 85 at week 4 and 80 at week 8. 84% would use the app again in the future. Median PrEP adherence was 91%. |
| Mitchell 2018<br>(USA)[38] | PrEP adherence monitoring | Descriptive quantitative | N = 12<br>Aged between 18-30 | Evaluation of the feasibility and acceptability of mSMART for PrEP – real-time camera-based adherence monitoring. Adherence increased for 30% of participants and remained stable for the other 70% who already had sufficient adherence. |

| **Study**<br>**(Location)** | **PrEP-related elements of**<br>**care delivered online** | **Design** | **Participants** | **Overview of study and main relevant findings** |
| --- | --- | --- | --- | --- |
| Refugio 2019<br>(USA)[39] | PrEP education; PrEP<br>eligibility assessment;<br>PrEP adherence support | Non-randomised<br>quantitative | N = 25<br>GBMSM<br>Median age = 22 | Evaluation of the feasibility of PrEPTECH where users create an account, test for HIV in at a lab, then receive 90-days' worth of PrEP. Phone consultations were conducted at days 30, 90 and 180. 85% of users said that PrEPTECH was a better way for GBMSM to access PrEP. |
| Siegler 2019<br>(USA)[40] | HIV testing (instructional<br>video); PrEP eligibility<br>assessment | Descriptive<br>quantitative | N = 58<br>Age: <30 = 38%;<br>30-39 = 40%;<br>40+ = 22% | Evaluation of PrEP@HOME – a service where participants completed self-sampling for HIV at home and an online behavioural survey resulting in a PrEP prescription being issued. Four participants required standard care. 93% were able to have their prescriptions renewed via PrEP@HOME. SUS score was 76.9. 85% said they would opt for PrEP@HOME over standard care. |
| Stekler 2018<br>(USA)[41] | PrEP education | Non-randomised<br>quantitative | N = 48 | Evaluation of the use of video calls within PrEP initiation appointments. No statistically significant difference between the proportion of participants prescribed PrEP within video call appointments compared to standard appointments. |
| Sullivan 2017<br>(USA)[42] | HIV testing (test ordering);<br>PrEP eligibility assessment<br>(including HIV risk<br>assessment) | Descriptive<br>quantitative | N = 121 | Assessment of the usability and acceptability of HealthMindr – an app with an HIV risk assessment, PrEP screening, HIV test comparison tool and HIV test ordering. Over 80% of users ordered a HIV test or condoms. 41% of those who ordered kits did so more than once. SUS score was 73.4. |

Seven studies described somewhat complete PrEP pathways.[29, 34-36, 39-41] Online modalities were employed in conjunction with face-to-face and telephone-based care and studies tended to rely on clinic or lab-based testing for HIV, renal function, and STIs.

#### Services focused on HIV testing without PrEP

Thirty-eight studies focused on HIV testing without explicitly being linked to PrEP. Seventeen studies (44.7%) allowed participants to order HIVSTs or self-sampling kits online, ten studies (26.3%) allowed patients to inform their healthcare providers of HIV test results online, and 16 (42%) allowed providers to inform patients of their results online. Participants were able to book face-to-face appointments for HIV testing online in two studies (5.3%), and five studies (13.2%) offered online HIV counselling. Eleven studies (29.0%) had an online HIV risk assessment. Some studies delivered multiple aspects of HIV-testing-related care online which accounts for the number of aspects of care exceeding the number of studies. However, 29 (76.3%) of the studies only delivered a single aspect of care online – mainly providers informing patients of their HIV test results (n=12, 41.4%) and HIVST or self-sampling kit ordering (n=9, 31.0%). An overview of these studies is found in Table 2.

**Table 2.** Overview of studies that delivered aspects of HIV testing not explicitly linked to PrEP.

| Study (Location) | Aspects of HIV testing delivered online | Design | Participants | Overview of study and summary of main findings |
| --- | --- | --- | --- | --- |
| Anand 2017b (Thailand)[43] | HIV counselling; ordering tests; booking face-to-face appointments; user to provider results; provider to user results | Non-randomised quantitative | N = 489<br>GBMSM and trans women<br>Median ages:<br>25 [23,28] (online arm);<br>29 [25,35] (hybrid arm);<br>27 [23,32] (offline arm) | Evaluation of myhealth.adamslove.org (electronic health records platform).<br>HIV diagnoses were highest among participants in the online arm (16%) compared to hybrid (1.4%) and offline arms (2.2%). |
| Balán 2020 (USA)[27] | User to provider results | Qualitative interviews and focus groups; descriptive quantitative | N = 59<br>GBMSM and trans women<br>Mean age (SD) = 40.8 (13.3) | Outline of users' preferences for a smartphone app that aimed to alleviate barriers associated with HIV self-tests. Users found the app acceptable and were able to use the app as intended during pilot testing. |
| Balán 2021 (USA)[44] | User to provider results | Descriptive quantitative; qualitative interviews | N = 48<br>GBMSM and trans women<br>Mean age (SD) = 41.7 (10.9) | Description of participants' use of the INSTI Multiplex and SMARTtest smartphone app. 77% of participants used the INSTI kit for self-tests and 54% used the kit for testing their partner(s). |
| Baraitser 2019 (England and Wales, UK)[45] | Ordering tests; user to provider results; HIV risk assessment | Descriptive quantitative | N = 1466<br>Mean age of self-test group = 28.6<br>Mean age of self-sample group = 27.0 | Evaluation of HIV self-testing and –sampling through SH24 – an online sexual health service. 67.1% of kits requested were self-tests (57.2% uploaded results); 32.9% were self-samples (53.9% of which were returned and results known). |

| **Study (Location)** | **Aspects of HIV testing delivered online** | **Design** | **Participants** | **Overview of study and summary of main findings** |
| --- | --- | --- | --- | --- |
| Bauermeister 2015 (USA)[46] | HIV risk assessment | Randomised controlled trial | N = 130<br>GBMSM<br>Median age = 21 | Evaluation of an online intervention that allowed user to identify HIV testing locations based on their specific structural needs.<br>31% of participants reported booking an appointment for testing. |
| Biello 2021b (USA)[47] | Ordering tests; provider to user results | Descriptive data from a randomised controlled trial | N = 80<br>GBMSM<br>Mean age (SD) = 20.7 (2.4) | Evaluation of the provision of HIV self-test and STI self-sample kits via the LYNX and MyChoices apps. Participants are emailed results if negative. 76% of those who completed the follow-up had tested for HIV during the study period, 54% reported using at least one HIV self-test. Acceptability was very high. |
| Chan 2021 (China)[48] | Ordering tests; pre- and post-test counselling | Descriptive quantitative | N = 350<br>GBMSM<br>Age: 13-30 (57.1%);<br>31-40 (31.4%);<br>>40 (11.4%) | Evaluation of an online service where people received a free self-test and booked a video call appointment via social media or phone call. 40.4% of new users and 63.1% of existing users ordered an test during the study period. 58.6% of new users and 79.4% of existing users tested for HIV during the study period. |
| Cohen 2017 (USA)[49] | Provider to user results | Non-randomised quantitative | N = 1460 | Evaluation of Healthvana, an online portal for accessing HIV and STI test results. Mean days between test and treatment was 10.68 for those who used Healthvana compared to 12.55 for those who did not. Mean days between test and notification of results was 6.86 for those who used Healthvana (8.77 for those who did not). |
| Daniels 2016 (USA)[50] | User to provider results | Descriptive quantitative | N = 37 | Evaluation of the acceptability of users sending photos of completed HIV self-tests to providers. Email was the preferred method of sending results by 37.8% of participants. |

| **Study**<br>**(Location)** | **Aspects of HIV testing**<br>**delivered online** | **Design** | **Participants** | **Overview of study and summary of main findings** |
| --- | --- | --- | --- | --- |
| De Boni 2019<br>(Brazil)[51] | Ordering tests | Descriptive<br>quantitative | N = 4800<br>GBMSM<br>Median age = 25 | Evaluation of a free, anonymous, internet-based HIV self-test strategy. Results were reported for 21.5% of the ordered tests. The study fulfilled all but one feasibility outcome measure. |
| Gilbert 2017<br>(Canada)[52] | Risk assessment; provider<br>to user results | Descriptive<br>quantitative | N = 868<br>Median age = 32 | Evaluation of GetCheckedOnline – a service where users complete an online assessment that suggests which STI tests should be completed at a lab (including HIV). Negative results were accessed online. 695 assessments were completed by 506 users. 12.5% were first time testers. |
| Gilbert 2018<br>(Canada)[53] | Risk assessment; provider<br>to user results | Descriptive<br>quantitative | N = 394<br>Median age:<br>GetCheckedOnline = 35;<br>Clinic = 30 | Evaluation of GetCheckedOnline – see Gilbert 2017. Compared to regular clinic attendees, GetCheckedOnline users were more likely to be older, test more frequently, and delay testing due to clinic distance and waiting times. |
| Gilbert 2019b<br>(Canada)[25] | Risk assessment; provider<br>to user results | Non-randomised<br>quantitative | N = 19,497 | Evaluation of GetCheckedOnline – see Gilbert 2017. GetCheckedOnline users were more likely to repeat test than regular clinic attendees. GetCheckedOnline users were more likely to be White, female, men who have sex with women, and a previous STI. |
| He 2018<br>(China)[54] | Provider to user results | Descriptive<br>quantitative | N = 500 | Evaluation of a service where people were offered anonymous HIV urine tests distributed through pharmacies. Participants accessed their results online via a unique code. 63% of the kits distributed were returned for analysis, 16.3% were reactive. 94.3% of people with reactive tests accessed their results; 83.7% of people with non-reactive tests accessed their results. |

| **Study (Location)** | **Aspects of HIV testing delivered online** | **Design** | **Participants** | **Overview of study and summary of main findings** |
| --- | --- | --- | --- | --- |
| He 2019 (China)[55] | Provider to user results | Descriptive quantitative | N = 957 | Evaluation of a service where people were offered anonymous HIV urine tests distributed by vending machines on university campuses. Participants access their results online via a unique code. 378 of 957 tests were returned; 376 were valid for analysis; 65.9% of people who tested negative accessed their results, 100% of people whose test was reactive accessed their results. |
| Hottes 2012 (Canada)[56] | Risk assessment; provider to user results | Focus groups | N = 39<br>Age: 20-29 (26%);<br>30-39 (13%);<br>40-49 (36%);<br>50+ (26%) | Exploration of the acceptability of internet-based STI and HIV testing where users complete an online assessment that suggests necessary tests which is taken to a lab. Results are accessed online. Participants were concerned about providing information online, other people entering their details, and how to interpret the tests. |
| Huang 2016 (USA)[57] | Ordering tests | Descriptive quantitative | N = 122<br>Age: 18-30 (65%); 31-40 (25%); 41+ (10%) | Evaluation of an HIV testing program linked to social media advertisements. 66.4% of participants ordered a test. Acceptability was high. |
| Jackman 2018 (USA)[58] | Provider to user results | Focus groups; descriptive quantitative | N = 35 (focus groups);<br>380 (survey) | Exploration of the use of electronic health records as a means of sexual health communication between partners. Participants thought that the records could help facilitate communication, particularly at the beginning of a relationship. |
| Jin 2018 (China)[59] | Risk assessment; order tests; user to provider results | Descriptive quantitative | N = 879<br>Median age = 28 | Assessment of the feasibility of Easy Test – an internet-based model for HIV self-testing. 40% of testers reported never having previously tested for HIV. 14.3% of tests were reactive. |

| **Study**<br>**(Location)** | **Aspects of HIV testing**<br>**delivered online** | **Design** | **Participants** | **Overview of study and summary of main findings** |
| --- | --- | --- | --- | --- |
| Knight 2019<br>(Canada)[60] | Risk assessment; provider to user results | Semi-structured interviews | N = 37<br>Mean age = 37.9 | Exploration of users' experiences with GetCheckedOnline and regular clinic-based care – see Gilbert 2017. Participants preferred GetCheckedOnline as it was more convenient and quicker, it provided them with more autonomy and control, and reduced their worries about being judged. |
| Lessard 2019<br>(France)[61] | Provider to user results; provider to user's partner(s) results notification | Focus groups | N = 21<br>Mean age = 38 | Exploration of the acceptability of WeFLASH© - a smartphone app where sexual partners link accounts, allowing providers to contact all relevant sexual partners immediately if someone tests positive for HIV or an STI. Acceptability was high. |
| MacGowan 2019<br>(USA)[62] | Ordering tests; user to provider results | Randomised controlled trial | N = 2665<br>GBMSM<br>Age: intervention group (18-29 (56.8%); 30+ (43.2%)); control group (18-29 (57.8%); 30+ (42.2%)) | Assessed the effect of providing HIV self-test kits on HIV testing. Participants in the intervention arm ordered and reported the results of the HIV self-tests on the study website. A higher proportion of participants in the intervention arm reported testing 3 or more times during the trial (76.6% vs 22.0%, $p < 0.01$). In the first three months, 0.9% of participants in the intervention arm were diagnosed with HIV compared to 0.1% of controls ($p < 0.01$). |
| Maksut 2014<br>(USA)[63] | Counselling | Descriptive quantitative | N = 20<br>Mean age = 28.1 | Piloted a video call-based HIV self-test support service. All participants found the service satisfactory and would use it again. |

| **Study (Location)** | **Aspects of HIV testing delivered online** | **Design** | **Participants** | **Overview of study and summary of main findings** |
| --- | --- | --- | --- | --- |
| Manavi 2017 (England, UK)[64] | Risk assessment; ordering tests | Descriptive quantitative | N = 5301<br>Median age = 24 | Evaluated Umbrella Health – a service that allows people to complete an assessment that determines what tests are needed. Women were more likely to return kits than men (61.2% v 53.1%, $p < .001$). GBMSM (62.5%) and women had a similar return rate (61.3% for non-anorectal, 59.6% for anorectal included). People preferred to have kits sent to their home (77.49% rather than 11.92% at clinic and 10.58% at pharmacy). |
| Menza 2021 (USA)[65] | Ordering tests | Descriptive quantitative | N = 233<br>Median age = 32 | Described the roll-out of Oregon’s state-wide pilot HIV self-test program. 233 people ordered 248 self-test kits. 73% of the initial 150 kits were ordered within the first 24 hours. 22% of participants lived in rural zip codes. |
| Page 2019 (England, UK)[66] | Ordering tests; provider to user results | Non-randomised quantitative | N = 550<br>Median age = 26 | Compared the feasibility of self-sampling for HIV using dry blood spot and micro-tube kits. 68.7% of MTs were returned compared to 66.5% of the DBS. 98.8% of DBS were successfully processed compared to 55.7% of the MTs ($p < 0.001$). |
| Phanuphach 2018 (Thailand)[67] | Counselling; ordering tests; appointment booking; user to provider results; provider to user results | Non-randomised quantitative | N = 571<br>GBMSM and trans women | Evaluated the acceptability and feasibility of myhealth.adamslove.org – an online HIV testing and linkage service. Choice of opting for the different aspects of the online service were associated with gender identity, income, social media and internet use, and past testing behaviours. |

| **Study**<br>**(Location)** | **Aspects of HIV testing**<br>**delivered online** | **Design** | **Participants** | **Overview of study and summary of main findings** |
| --- | --- | --- | --- | --- |
| Polilli 2016<br>(Italy)[68] | Risk assessment;<br>appointment booking | Descriptive<br>quantitative | N = “about 6000” | Evaluation of a web-based HIV testing initiative where participants could complete an online HIV risk assessment then book an in-person appointment. 28 (0.92%) of the people tested were diagnosed with HIV; 26 (92.8%) were linked to care. |
| Rosengren<br>2016<br>(USA)[69] | Ordering tests | Descriptive<br>quantitative | N = 125<br>GBMSM<br>Age: 18-30 (63%); 31-40<br>(30%); 41+ (8%) | Evaluated the distribution of HIV self-test kits through Grindr. 59% said self-testing was very easy and 34% said it was easy. When comparing the HIVST kit and clinic testing, 35 (63%) preferred self-testing kit and 8 (14%) somewhat preferred self-testing. |
| Salway 2019<br>(Canada)[70] | Risk assessment; provider<br>to user results | Non-randomised<br>quantitative | N = 352<br>Median age = 35 (GCO);<br>30 (regular clinic) | Compared GetCheckedOnline users’ (see Gilbert 2017) HIV test knowledge and sexual behaviours with users accessing regular clinics. GetCheckedOnline users showed higher HIV knowledge post-test at baseline. HIV knowledge was associated with Canadian residency, education, use of English, and being GBMSM. |
| Stephenson<br>2020<br>(USA)[71] | Counselling; user to<br>provider results | Randomised<br>controlled trial | N = 202<br>Binary and non-binary<br>transgender youth | Evaluated Project Moxie – a service where participants attend a video call appointment with an HIV counsellor who provides real-time support for people while they complete an HIV self-test. The self-test was performed in 96% of video call sessions and satisfaction was very high. |
| Syred 2019<br>(England,<br>UK)[72] | Ordering tests | Non-randomised<br>quantitative | N = 6254 | Described a ‘choose to test’ service where participants completed an assessment that then recommends what tests they should order. 7550 tests were ordered by 6253 users. People chose to add and remove tests. |

| **Study (Location)** | **Aspects of HIV testing delivered online** | **Design** | **Participants** | **Overview of study and summary of main findings** |
| --- | --- | --- | --- | --- |
| Wang 2018 (China)[73] | Counselling | Randomised controlled trial | N = 430<br>Age: 18-30 (63.1%); 31+ (36.9%) | Evaluation of the efficacy of real-time instructions and counselling for HIV self-testing delivered via video call. HIV testing was higher in the intervention group than the control group at 5 months (89.8% v 50.7%; $p < .001$). Self-reported uptake of HIVST was 87.9% in the intervention arm compared to 2.3% in controls ($p < .001$). |
| Wilson 2017 (England, UK)[74] | Ordering tests | Randomised controlled trial | N = 2072<br>Mean age = 23 | Evaluation of the effect of SH24 (an online STI testing and results service) on uptake of STI testing (including HIV). 50% of intervention group and 26.6% of the control group had completed an STI test by week 6 (RR 1.87, $p < .001$). |
| Witzel 2019 (England and Wales, UK)[26] | Ordering tests | Randomised controlled trial; focus groups | N = 1035 (RCT); 10 (focus group)<br>Median age = 32.1 | Evaluation of the feasibility of an online recruitment strategy for an RCT looking at online provision of HIV self-test kits. The strategy appeared feasible and acceptable. Participants noted several barriers including the blood-borne nature of the tests and anxiety. |
| Witzel 2021 (England and Wales, UK)[75] | Ordering tests | Randomised controlled trial; semi-structured interviews | N = 295<br>Median age = 29 | Evaluation of online provision of HIV self-tests focusing on trans participants. In trans men, HIV testing uptake was significantly higher in the baseline test group (95%) than in the no baseline test group (29%) ($p < 0.001$). Acceptability was very high. Perceived benefits included: increased autonomy, privacy, convenience and avoidance of discrimination and dysphoria. |
| Wray 2018 (USA)[76] | Notification of test initiation | Randomised controlled trial | N = 65<br>Mean age = control (35.5); standard (35.8); eTEST (34.1). | Evaluation of the use of beacons that signal when HIV self-tests are opened, allowing providers to follow-up and offer support. All participants in the arms that provided HIV self-tests tested for HIV within the study period. |

| **Study**<br>**(Location)** | **Aspects of HIV testing**<br>**delivered online** | **Design** | **Participants** | **Overview of study and summary of main findings** |
| --- | --- | --- | --- | --- |
| Xia 2018<br>(China)[77] | Provider to user results | Descriptive<br>quantitative | N = 3092 | Evaluated anonymous urine testing for HIV (see X). 3092 kits were distributed, 1977 kits were returned, 1911 were eligible for analysis, and 145 were reactive. 83% of people with reactive tests accessed their results online. |

#### Other studies

Only one study was related to renal function monitoring and was not linked to PrEP.[78] This study described a website-based patient portal where patients could access their renal test results.[78] Participants found the portal easy to use (92%) and said that it helped them manage their conditions (93%).

The reviews of online content included in this review all related to PrEP education.[79-82] Whiteley *et al*. evaluated websites and YouTube videos that provided PrEP information and found that no website fully satisfied their four appraisal criteria.[82] Kecojevic *et al*. evaluated YouTube videos that provided information on PrEP and found that the videos varied in terms of the completeness of the information provided.[81] Gilbert *et al*. evaluated the information relating to HIV risk and prevention on Canadian HIV websites (this included PrEP). They highlighted the potential accessibility challenges found in the websites (e.g. high reading level) and the low usability and lack of interactive features included in the websites.[79] Deviating slightly from the other reviews which focused on information readily available to users, Lee *et al*. conducted a retrospective analysis of questions submitted to an online HIV counselling website about HIV and PrEP – in this case, it was the service users requesting information about PrEP.[80] Questions tended to revolve around HIV testing, self-perceived HIV risk, emotional state, and treatment and prevention.

### Acceptability and feasibility of online PrEP-specific care

Acceptability was measured in 30 studies either qualitatively through focus groups and interviews (n=9) or in a cross-sectional survey (n=23). Full details on acceptability can be found in **Supplementary Material 5**. Overall, acceptability was very high. Participants praised the convenience of the online services and the added privacy they provided, as well as reporting a positive overall experience. Four studies used the System Usability Scale (SUS), a validated measure of subjective usability.[83] SUS scores ranged from 68.25 to 85;[24, 37, 38, 40] scores above 68 are considered above average in terms of usability.[83]

We extracted data on service uptake, retention and service delivery to summarise feasibility given the heterogeneity of the included studies – this can be found in **Supplementary Material 5**. The included studies appeared to be able to recruit and retain a sufficient sample to address their specific aims, and appeared to be able to deliver their services as intended. Notably, studies that focused on online HIV testing were able to recruit a high number of people who were either engaging in higher risk behaviours or who were unaware that they were already living with HIV.[28, 30, 43, 54, 59, 67, 77] Moreover, one study was able to demonstrate the feasibility of having patients self-sample for creatinine, with 81% of the self-samples being adequate for analysis.[29]

### Barriers to, and facilitators of, online PrEP-related care

Nineteen studies identified barriers, facilitators and other factors that influenced engagement with their specific service. Herein, we focus solely on the barriers, facilitators and factors linked to the PrEP-related elements of care that were delivered online.

Convenience was identified as a facilitator in eight studies;[26, 29, 35, 36, 56, 60, 63, 65] in particular, the ability of online services to help overcome geographic barriers and scheduling issues.[26, 29, 56, 60, 63] Participants cited the need to attend a specific physical location for testing as a barrier.[36] However, blood-based self-sampling was also reported as a barrier for people who had not performed the procedure before.[26]

Participants highlighted the importance of reminding patients that face-to-face care was still available,[56] and that online care should be seen as complementary to face-to-face care rather than a replacement.[60] Quarterly testing appeared to facilitate a routine for HIV and STI testing,[36] and notifications to remind participants to take PrEP were seen to facilitate service engagement.[38] Participants also felt that online testing may not be suitable for first time testers.[56] Academic language was reported as a barrier to comprehension and engagement with services.[24] Participants also highlighted the importance of striking a balance between efficiency and having a human touch within online service delivery.[35]

Privacy was a recurring theme across four studies. Most accounts suggested that the online nature of the services facilitated privacy.[26, 60] However, participants also highlighted that the perceived security and reliability of postal services affected their decision to opt for home or laboratory based screening (i.e. low perceived security/reliability would make home screening less appealing).[29] Participants also reported that they felt a reluctance to provide personal information online.[56]

Three studies indicated that having parts of the care pathway delivered online allowed participants to avoid stigma, judgement and embarrassment felt when accessing care face-to-face.[29, 56, 63] In contrast, participants had differing views on the impact of HIV self-tests on anxiety.[26] Some felt that using HIV self-tests ordered online increased anxiety levels because it was removed from traditional care and would leave them feel isolated when facing a positive result.[26] Conversely, those who were more experienced testers felt less anxiety because the HIVST gave results almost immediately.[26] Participants also felt that online care provided them with more autonomy and control and was perceived as less invasive than traditional care.[60]

## DISCUSSION

### Main findings

The 59 included articles varied in design, methodology, and the type and breadth of PrEP-related care provided. Online PrEP pathways, or at least parts of them, appeared broadly acceptable and feasible. Seven articles described complete PrEP care pathways that used a combination of online, face-to-face, and telephone-based care.[29, 34-36, 39-41] However, none described a service where a complete PrEP pathway was delivered online (i.e. online consultation, remote self-sampling, and an electronic PrEP prescription). Online care was primarily delivered through websites, video calls, and smartphone apps. The acceptability and feasibility of online PrEP care was promising; the included studies were able to recruit and retain participants and deliver their services as intended. Online services were able to reach key populations and identified people who were unaware they were living with HIV and link them to care.[28, 30, 43, 54, 59, 67, 77] Where measured, participants rated the acceptability and usability of services highly and the four studies that measured acceptability using the System Usability Scale each scored above average.[24, 37, 38, 40]

Most elements that would form part of a complete online PrEP pathway have been delivered online individually within the context of PrEP or in broader health contexts (e.g. HIV testing). HIV testing underpins safe PrEP provision because providing tenofovir and emtricitabine to people living with undiagnosed HIV could drive resistance to antiretroviral medication more broadly and compromise future treatment options.[10] It is therefore unsurprising that the majority of studies included one or more aspect of online HIV testing (e.g. ordering an HIV self-test or self-sampling kit, accessing results online). Renal function monitoring was only mentioned in three articles: one outlined an online portal where patients could access their renal test results,[78] one described a system where people could book a face-to-face renal function test online,[34] and one evaluated a service where patients could order self-sample kits for creatinine analysis.[29] The latter is particularly important in the context of remote or online PrEP care given that the estimated glomerular filtration rate (eGFR) is typically used as an indicator of renal health and uses serum creatinine levels in its calculation.[10] No study reported monitoring side effects and possible drug interactions online; however, it is possible that this was addressed in studies within PrEP eligibility assessments. This is important because side effects are likely to influence adherence to PrEP, which in turn determines efficacy.[84-86]

Online care appeared to help overcome some of the barriers people experienced when accessing face-to-face care. Participants found online services both convenient, and able to overcome geographical barriers.[26, 29, 56, 60, 63] Participants in qualitative studies also found that online care facilitated privacy and avoided the stigma sometimes felt when accessing services in person.[26, 29, 56, 60, 63] This was echoed in the quantitative data where online services were able to reach people who had never tested for HIV before.[28] While this is a strength of online care modalities, it also highlights the ongoing issue of stigma and the need for continued and enhanced efforts to reduce stigma in healthcare settings irrespective of mode of delivery.[87, 88] In contrast, online care appeared to introduce or exacerbate some barriers to access. Self-sampling for blood borne viruses was seen as a barrier where people had not performed the procedure before.[26, 56] Some participants also felt a reluctance to provide information online.[56]

### Comparison with existing literature

We are not aware of any other literature reviews that cover online PrEP care to date aside from a 2019 narrative overview of telehealth interventions aiding PrEP delivery.[89] Compared to this earlier review, which also concluded that telehealth interventions have the potential to strengthen the PrEP care continuum, we adopted a wider definition of PrEP care. Our definition was based on clinical recommendations for safe PrEP prescribing and we conducted our search over two years since the earlier review was published, which is particularly relevant given the shifts towards remote, digital health observed during the COVID-19 pandemic.[10, 90, 91]

### Strengths and weaknesses

We followed established guidelines for conducting and reporting scoping reviews.[15-17] Our choice of scoping rather than systematic review methodology meant that we were able to include a diverse range of published studies in an emerging field with few randomised controlled trials. The majority of included studies were non-randomised, descriptive or used qualitative methods which is unsurprising given the relative novelty of online PrEP care. However, this limited our ability to synthesise outcomes. The included articles did not consistently report their design. To remedy this and aid consistency across the review, we categorised the included studies using the design groups outlined in the Mixed Methods Appraisal Tool (e.g. randomised controlled trial, descriptive quantitative study, qualitative).[23] Moreover, we made a distinction between online (e.g. email, video call, internet-based instant messaging) and SMS/phone-based care for this review. On reflection, it is unclear if this would be a meaningful distinction from a patient perspective.

### Implications for practice and future research

None of the articles included described a service that delivered a complete PrEP pathway online (i.e. online consultation, remote self-sampling, and an electronic prescription). Although each of these stages appears feasible, delivered online with high acceptability; the overall feasibility and acceptability of combining these stages within one seamless online PrEP pathway remains unclear. The evidence collected in this review suggests that a complete online PrEP service could be possible; certainly, the feasibility of patients self-sampling for creatinine in the context of PrEP care is a promising indicator of the extent to which PrEP-related care could be self-managed. Further research is needed to develop and evaluate a complete online PrEP pathway, paying close attention to clinical outcomes. We should also be mindful of digital exclusion when designing services, ensuring that PrEP can be accessed in a variety of ways to meet the needs and preferences of key populations.[92] Increasing PrEP coverage in key populations may be instrumental in achieving the UNAIDS targets and online provision of PrEP care appears to be a promising, scalable addition to help achieve these goals.[12]

## Conclusions

Online provision of elements of PrEP care is feasible and acceptable to service users. Online PrEP care appears to be able to alleviate some of the barriers associated with current PrEP pathways; however, it also introduces, or exacerbates, other barriers. The integration of online care into PrEP provision has the potential to strengthen efforts to achieve the UNAIDS goals of zero HIV transmissions by appealing to key populations and providing economies of scale.[12] In order to understand the full potential of online PrEP provision, additional research is needed to understand the acceptability and feasibility of a seamless, online PrEP service that can integrate with existing services.

## Data Availability

All data produced in the present study are available upon reasonable request to the authors.

## FUNDING STATEMENT

This work is funded by NHS Greater Glasgow & Clyde, NHS Lothian, and Public Health Scotland.

## INTERESTS STATEMENT

None to declare.

## CONTRIBUTORS

RK, CE, JG, JF and JD contributed to the conception and design of the study. RK, JG and CE contributed to the establishment of the search strategy. RK and JG contributed to establishment of the method of analysis. RK led the article reviewing and data analysis with contribution from JG. RK drafted the original manuscript with guidance from CE, JF, JG and JD. All authors (RK, CE, JF, JD and JG) have made contributions to the drafting and revising of the article and have approved the final version.

## DATA AVAILABILITY STATEMENT

All data relevant to the study are included in the article and supplementary materials. This manuscript reports on a scoping review. All articles included are included in the article or in the supplementary materials.

## Notes

### Competing Interest Statement

The authors have declared no competing interest.

### Clinical Protocols

https://doi.org/10.6084/m9.figshare.17157899

