## Supplementary Material 1 for "Delivering HIV pre-exposure prophylaxis (PrEP) care online: A scoping review"

**Supplementary Material 1 – Defining PrEP Care and Search Strategy**

An HIV pre-exposure prophylaxis (PrEP) prescription, like any other medication, is usually prescribed after it is deemed safe and appropriate for that individual. This typically involves assessment of HIV risk and medical tests every three months. [1] Given that PrEP is provided alongside other interventions, focusing solely on studies that explicitly prescribed PrEP would have fell short of the aim of this review – to understand the extent to which PrEP care had been delivered online. The PrEP prescription, while the central aspect of PrEP care, is only one facet. Therefore, we developed a comprehensive definition of PrEP care on which to base this review.

**Translating recommendations to behaviours**

The British HIV Association and British Association for Sexual Health and HIV joint recommendations for PrEP use were used as the basis for developing the definition of PrEP care. [1] The recommendations were chosen as the basis of developing the definition as they were evidence-based and comprehensive. While the completeness of the recommendations was critical, this did not lend itself to easily forming a search strategy, hence the need to rework the recommendations into a usable definition of PrEP care.

The review forms part of a larger research project which adopts a behavioural approach to intervention development. Therefore, the first step in developing the definition was to translate the recommendations into behaviours. This is not particularly relevant to this review; however, it felt important to note for completeness.

**Grouping behaviours into components**

Once the recommendations were translated into behaviours and reviewed by the wider study team, the behaviours were grouped based on the similarity of care they provided; for example, all behaviours relating to PrEP adherence were grouped. This resulted in 12 groups, or elements of PrEP-related care: HIV testing; renal monitoring; PrEP prescription; PrEP eligibility assessment/HIV risk assessment; PrEP adherence monitoring and support; PrEP education; PrEP-related side-effects and drug interactions; post-exposure prophylaxis; testing for sexually transmitted infections; bone mineral density assessment; hepatitis testing and care; and vaccinations.

**Focusing on essential elements of PrEP care**

When considering the elements of PrEP care, there appeared to be a great deal of heterogeneity in type of care and the necessity of that care for safe PrEP prescription. We therefore decided to focus on the elements of care that were deemed to be essential to safe and appropriate PrEP prescription. Our review focused on the following elements of PrEP-related care: HIV testing; renal monitoring; PrEP prescription; PrEP eligibility assessment; PrEP education; PrEP adherence monitoring and support; and PrEP-related side-effects and drug interactions.

**Search strategy**

The aim of this review was to explore the extent to which PrEP-related care had been delivered online. As we were interested in understanding the scope of PrEP-related elements of care that were not necessarily conducted in the context of PrEP (e.g. HIV testing), simply searching for PrEP and its variations (e.g. preexposure prophylaxis) would not have been sufficient to meet the aims. This lead to a somewhat complex search consisting of four independent searches: HIV testing, renal monitoring, PrEP prescription, and general PrEP care. Table 1 shows how the PrEP-specific care components mapped onto these searches. A general search for PrEP care covered most components because individual searches would have used the same terms but with additional terms that would have further limited the scope of the search. A wider search, therefore, would identify any relevant articles that multiple narrower searches would have and is also in keeping with the scoping nature of this review. The PrEP prescription search is separate because of the way in which the terms were combined. This is explained in more detail below.

**Table 1.** *Mapping of elements of care onto searches.*

| **Search** | **PrEP-specific elements of care** |
| --- | --- |
| HIV testing | HIV testing |
| Renal monitoring | Renal Monitoring |
| PrEP prescription | PrEP prescription |
| General PrEP search | PrEP adherence monitoring and support |
|  | PrEP eligibility assessment |
|  | PrEP education |
|  | PrEP side effects and drug interactions |

**PrEP terms**

The PrEP search terms were developed by combining the various ways in which PrEP tends to be referred to in the literature. Through trialling the different combinations of terms we used the following terms: “PrEP” OR “pre-exposure prophylaxis” OR “preexposure prophylaxis” OR “pre exposure prophylaxis”. Where available, the MeSH term “pre-exposure prophylaxis” was also included, added with the “OR” Boolean operator.

**Online terms**

Search terms for online care were adapted from an existing systematic review of digital innovations for STIs and HIV and were as follows: “telemedicine” OR “eHealth” OR “e-health” OR “mHealth” OR “m-health” OR “mobile health” OR “mobile technology” OR “mobile applications” OR “app” OR “apps” OR “social medi*” OR “cell phone*” OR “cellphone*” OR “mobile phone*” OR “mobile telephone*” OR “cellular phone*” OR “smartphone*” OR “smart phone*” OR “mobile device*” OR “text messag*” OR “texting” OR “texted” OR “SMS” OR “MMS” OR “multimedia messag*” OR “short messag*” OR “computers, handheld” OR “personal digital assistant” OR “email*” OR “e-mail*” OR “online” OR “internet” OR “web” OR “digital health” OR “remote*”. [2] Where available, the MeSH terms “telemedicine” and “mobile applications” were also included, added with the “OR” Boolean operator. In hindsight, we could have paired this down to remove SMS-related terms given that studies that focused on SMS were not included; however, at the time we decided to keep the terms for a more complete search.

**Terms for PrEP care components**

Search terms for HIV testing were adapted from an existing systematic review of HIV testing within general practice. [3] We felt that adding the “*” operator would provide a more comprehensive search by allowing for variations of test and screen (e.g. tests and testing). This resulted in the following terms: “HIV test*” OR “HIV screen*” OR (“HIV” AND “test*”) OR (“HIV” AND “screen*”).

The renal monitoring search terms were adapted from a systematic review of renal health in healthy individuals. [4] The search terms were: “kidney function test*” OR “renal function*” OR “creatinine” OR “proteinuria” OR “urinalysis”.

The online prescription search terms incorporated the online care search terms outlined above. The online care search terms were combined with “prescri*” and combined with the MeSH terms ‘drug therapy, computer assisted’, ‘electronic prescribing’, ‘medical order entry system’ and ‘pharmaceutical services, online’ where available. This resulted in the following search: (***online care terms*** AND “prescri*”) OR “drug therapy, computer assisted” OR “electronic prescribing” OR “medical order entry system” OR “pharmaceutical services, online”.

**Combining search terms**

Once terms were identified for PrEP, online care, and each of the PrEP-specific elements of care, the four searches were created by various combinations of these terms. For HIV testing and renal monitoring, terms specific to those elements of care were combined with the terms for online care using the AND Boolean operator. The PrEP terms were not included in these searches as we were interested in any online HIV testing or renal monitoring, not just that which was delivered in conjunction with PrEP. Because we chose to use a general PrEP search to capture many of the care components, we did not need to identify terms specific to these components. Instead, we combined the PrEP terms and the online care terms using the AND Boolean operator. PrEP prescription was run as a separate search because the specific terms for online prescription already included the online terms.

**Table 2.** *Combining search terms.*

|  | **Terms specifically for that component?** | **AND PrEP terms?** | **AND online terms?** |
| --- | --- | --- | --- |
| **HIV testing** | Yes | No | Yes |
| **Renal monitoring** | Yes | No | Yes |
| **PrEP prescription** | Yes | Yes | No |
| **General PrEP search** | No | Yes | Yes |
