## Supplementary Material 3 for "Delivering HIV pre-exposure prophylaxis (PrEP) care online: A scoping review"

**Supplementary Material 3 - Mixed Methods Appraisal Tool Overview**

In line with the recommended procedure for MMAT, if the article did not fulfil the initial screening questions, it was deemed low quality and further appraisal was not performed.

**MMAT summary for quantitative randomized controlled trials**

| **Study ID** | **Screening questions** | | **Quantitative randomized controlled trial-specific questions** | | | | |
| --- | --- | --- | --- | --- | --- | --- | --- |
|  | **Are there clear research questions?** | **Do the collected data allow to address the research questions?** | **Is the randomization appropriately performed?** | **Are the groups compared at baseline?** | **Are there complete outcome data?** | **Are outcome assessors blinded to the intervention provided?** | **Did the participants adhere to the assigned intervention?** |
| Bauermeister 2015 | Yes | Yes | Unclear | No | No | Unclear | Yes |
| MacGowan 2019 | Yes | Yes | Yes | Yes | No | Unclear | Unclear |
| Stephenson 2020 | Yes | Yes | Yes | Yes | Unclear | Unclear | Unclear |
| Wang 2018 | Yes | Yes | Yes | Yes | Unclear | No | Unclear |
| Wilson 2017 | Yes | Yes | Yes | Yes | Unclear | Yes | Unclear |
| Witzel 2019 | Yes | Yes | Yes | Yes | Unclear | Unclear | Unclear |
| Witzel 2021 | Yes | Yes | Yes | Yes | Unclear | No | Unclear |
| Wray 2018 | Yes | Yes | Unclear | Yes | Unclear | No | Unclear |

**MMAT summary for quantitative non-randomised studies**

| **Study ID** | **Screening questions** | | **Quantitative non-randomized study-specific questions** | | | | |
| --- | --- | --- | --- | --- | --- | --- | --- |
|  | **Are there clear research questions?** | **Do the collected data allow to address the research questions?** | **Are the participants representative of the target population?** | **Are the measurements appropriate regarding both the outcome and intervention (or exposure)?** | **Are there complete outcome data?** | **Are the confounders accounted for in the design analysis?** | **During the study period, is the intervention administered (or exposure occurred) as intended?** |
| Anand 2017a | Yes | Yes | Unclear | Yes | No | No | Yes |
| Anand 2017b | Yes | Yes | Unclear | Yes | No | Yes | Yes |
| Chasco 2021 | Yes | Yes | Unclear | Yes | Yes | Unclear | Yes |
| Cohen 2017 | Yes | Yes | Yes | Yes | Yes | Unclear | Yes |
| Fuchs 2018 | Yes | Yes | Unclear | Yes | No | Unclear | Yes |
| Gilbert 2019b | Yes | Yes | Yes | Yes | Yes | Yes | Yes |
| Page 2019 | Yes | Yes | Unclear | Yes | No | Unclear | Unclear |
| Phanuphack 2018 | Yes | Yes | Unclear | Yes | Unclear | Yes | Unclear |
| Refugio 2019 | Yes | Yes | Unclear | Yes | Unclear | Unclear | Yes |
| Salway 2019 | Yes | Yes | Unclear | Yes | Unclear | Yes | Unclear |
| Stekler 2018 | Yes | Yes | Unclear | Yes | No | Unclear | Yes |
| Syred 2019 | Yes | Yes | Yes | Yes | Yes | Unclear | Yes |

**MMAT summary for quantitative descriptive studies**

| **Study ID** | **Screening questions** | | **Quantitative descriptive study-specific questions** | | | | |
| --- | --- | --- | --- | --- | --- | --- | --- |
|  | **Are there clear research questions?** | **Do the collected data allow to address the research questions?** | **Is the sampling strategy relevant to address the research question?** | **Is the sample representative of the total population?** | **Are the measurements appropriate?** | **Is the risk of nonresponse bias low?** | **Is the statistical analysis appropriate to answer the research question?** |
| Balán 2020 | Yes | Yes | Unclear | Unclear | Unclear | Unclear | Unclear |
| Balán 2021 | Yes | Yes | Yes | Unclear | Yes | Yes | Yes |
| Baraitser 2019 | Yes | Yes | Yes | Unclear | Yes | No | Yes |
| Biello 2021a | Yes | Yes | Yes | Unclear | Yes | Unclear | Yes |
| Biello 2021b | Yes | Yes | Yes | Unclear | Yes | Unclear | Yes |
| Chan 2021 | Yes | Yes | Unclear | Unclear | Yes | No | Yes |
| Daniels 2016 | Yes | Yes | Yes | Unclear | Yes | Unclear | Yes |
| De Boni 2019 | Yes | Yes | Yes | Unclear | Yes | No | Yes |
| Elliot 2016 | Yes | Yes | Yes | Unclear | Yes | Unclear | Yes |
| Finkenflügel 2019 | Yes | Yes | Yes | Unclear | Yes | Yes | Yes |
| Gilbert 2017 | Yes | Yes | Yes | Yes | Yes | Unclear | Yes |
| Gilbert 2018 | Yes | Yes | Yes | Unclear | Yes | Unclear | Yes |
| Guinness 2018 | Yes | Yes | Yes | Yes | Yes | Unclear | Yes |
| He 2018 | Yes | Yes | Unclear | Unclear | Unclear | No | Unclear |
| He 2019 | Yes | No | N/A | N/A | N/A | N/A | N/A |
| Hoth 2019 | Yes | Yes | Yes | Yes | Yes | No | Yes |
| Huang 2016 | Yes | Yes | Yes | Unclear | Yes | No | Yes |
| Jackman 2018 | Yes | Yes | Yes | Unclear | Yes | No | Yes |
| Jin 2018 | Yes | Yes | Yes | Unclear | Yes | No | Yes |
| Liu 2021 | Yes | Yes | Unclear | Unclear | Yes | Yes | Yes |
| Maksut 2016 | Yes | Yes | Yes | Yes | Yes | Unclear | Yes |
| Manavi 2017 | Yes | Yes | Yes | Yes | Yes | Unclear | Yes |
| Mitchell 2018 | Yes | Yes | Yes | Unclear | Yes | Unclear | Yes |
| Menza 2021 | Yes | Yes | Yes | Unclear | Yes | Unclear | Yes |
| Polilli 2016 | Yes | Yes | Yes | Unclear | Unclear | Unclear | Unclear |
| Rosengren 2016 | Yes | Yes | Yes | Unclear | Unclear | Unclear | Yes |
| Siegler 2019 | Yes | Yes | Yes | Unclear | Yes | Unclear | Yes |
| Sullivan 2017 | Yes | Yes | Yes | Unclear | Yes | Unclear | Yes |
| Woywodt 2014 | Yes | Yes | Yes | Yes | Yes | No | Yes |
| Xia 2018 | Yes | Yes | Yes | Unclear | Yes | Unclear | Yes |

**MMAT summary for qualitative studies**

| **Study ID** | **Screening questions** | | **Qualitative study-specific questions** | | | | |
| --- | --- | --- | --- | --- | --- | --- | --- |
|  | **Are there clear research questions?** | **Do the collected data allow to address the research questions?** | **Is the qualitative approach appropriate to answer the research question?** | **Are the qualitative data collection methods adequate to address the research question?** | **Are the findings adequately derived from the data?** | **Is the interpretation of results sufficiently substantiated by data?** | **Is there coherence between qualitative data sources, collection, analysis and interpretation?** |
| Balán 2020 | Yes | Yes | Yes | Yes | Yes | Yes | Yes |
| Balán 2021 | Yes | Yes | Yes | Yes | Yes | Yes | Yes |
| Biello 2021a | Yes | Yes | Yes | Yes | Yes | Yes | Yes |
| Chasco 2021 | Yes | Yes | Yes | Yes | Yes | Yes | Yes |
| Fuchs 2018 | Yes | Yes | Yes | Yes | Yes | Yes | Yes |
| Hottes 2012 | Yes | Yes | Yes | Yes | Yes | Yes | Yes |
| Hughes 2021 | Yes | Yes | Yes | Yes | Yes | Yes | Yes |
| Jackman 2018 | Yes | Yes | Yes | Yes | Yes | Yes | Yes |
| Knight 2019 | Yes | Yes | Yes | Yes | Yes | Yes | Yes |
| Koester 2020 | Yes | Yes | Yes | Yes | Yes | Yes | Yes |
| Lessard 2019 | Yes | Yes | Yes | Yes | Yes | Yes | Yes |
| Liu 2021 | Yes | Yes | Yes | Yes | Yes | Unclear | Unclear |
| Witzel 2019 | Yes | Yes | Yes | Yes | Yes | Yes | Yes |
| Witzel 2021 | Yes | Yes | Yes | Yes | Yes | Yes | Yes |

**MMAT summary for mixed methods studies**

Note: quality appraisal was conducted for the specific methods used in these mixed methods studies in the preceding tables

| **Study ID** | **MMAT methodology categories used** | **Mixed methods-specific questions** | | | | |
| --- | --- | --- | --- | --- | --- | --- |
|  |  | **Is there an adequate rationale for using a mixed methods design to address the research question?** | **Are the different components of the study effectively integrated to answer the research question?** | **Are the outputs of the integration of qualitative and quantitative components adequately interpreted?** | **Are divergences and inconsistencies between qualitative and quantitative research adequately addressed?** | **Do the different components of the study adhere to the quality criteria of each tradition of the methods involved?** |
| Balán 2020 | Quantitative descriptive and qualitative | Yes | Yes | Unclear | N/A | No |
| Balán 2021 | Quantitative descriptive and qualitative | Yes | Yes | Yes | N/A | Yes |
| Biello 2021a | Quantitative descriptive and qualitative | Yes | Yes | Yes | N/A | Yes |
| Chasco 2021 | Quantitative non-randomised and qualitative | Yes | Yes | Yes | N/A | Yes |
| Fuchs 2018 | Quantitative non-randomised and qualitative | Yes | Yes | Yes | N/A | Unclear |
| Jackman 2018 | Quantitative descriptive and qualitative | Yes | Yes | Yes | N/A | Unclear |
| Liu 2021 | Qualitative and descriptive quantitative | Yes | Yes | Yes | N/A | Unclear |
| Witzel 2019 | RCT and qualitative | Yes | Yes | Yes | N/A | Unclear |
| Witzel 2021 | RCT and qualitative | Yes | Yes | Yes | N/A | Unclear |
