## Supplementary Material 4 for "Delivering HIV pre-exposure prophylaxis (PrEP) care online: A scoping review"

| **Author(s)** | **Design, participants, location and study length** | **Overview of study/service** | **Summary of main findings** |
| --- | --- | --- | --- |
| **Anand 2017a** | Non-randomised quantitative (self-selecting trial)  N=489  **PrEP-C.C.:** HIV testing (counselling, self-test ordering, appointment booking, user to provider and provider to user results)  **Modality:** website, video call  **Location:** Thailand | Examination of factors associated with the uptake and use of the different pathways within myhealth.adamslove.org (electronic health records) for HIV testing: offline (face-to-face care and referred to the website for info post-care); online (supervised HIV self-testing entirely remotely with virtual care); and hybrid (online pre-test counselling and appointment booking, HIV test completed at clinic). | HIV prevalence was highest in the online arm (16%) compared to the hybrid and offline arms (1.4% and 2.2%, respectively).  Participants in the online arm were less likely to access lab results (33.3%) than in the offline and hybrid arms (79.1% and 71.1%, respectively).  Participants in the online arm were less likely to read their post-test summaries (33.3%) compared to the offline and hybrid arms (53.5% and 55.6%). |
| **Anand 2017b** | Non-randomised quantitative (self-selecting feasibility trial)  N=425  **PrEP-C.C.:** HIV testing (appointment booking); PrEP education; PrEP eligibility assessment (and HIV risk assessment)  **Modality:** website (including instant messaging), video call  **Location:** Thailand | The study evaluated Adam’s Love – an online to offline model for HIV testing and PrEP. Adam’s Love was reported to offer eCounselling, PrEP education, an HIV risk assessment, and online booking for in person clinical care. | Of the 425 people who received the online intervention, 325 (76.5%) checked in at the appointment.  PrEP uptake was highest among the Adam’s Love clinic (70.4%) compared to the Thai Red Cross AIDS Research Centre (46.7%) and community based clinics (39.1%). |
| **Balán 2020** | Qualitative interviews, focus groups and descriptive quantitative  N=59  **PrEP-C.C.:** HIV testing (user to provider results)  **Modality:** Smartphone app, email  **Location:** USA | The study describes users’ preferences for a smartphone app that aims to alleviate some of the barriers associated with HIV self-testing. It also described how this was integrated with another app (SMARTtest) which supports self- and partner-testing. The app allows users to send their test results to their doctor via email. | Participants found the function to share test results with their doctor highly acceptable but wanted to control how these results were stored and shared.  From pilot testing (n=9), all participants were able to scan their results but the accuracy of the interpretations made from these scanned tests varied. Some participants who received invalid or incorrect results were able to visually interpret their results which led to changes being made to the app. |
| **Balán 2021** | Descriptive quantitative (feasibility) and qualitative interviews  N=48  **PrEP-C.C.:** HIV testing (user to provider results)  **Modality:** Smartphone app, email  **Location:** USA | The study describes participants’ use of the INSTI Multiplex for HIV self- and partner testing. The app allows users to send their test results to their doctor via email. | Seventy-seven percent of participants self-tested using the INSTI kit (mean 3.7 times; SD = 3.9 times).  Fifty-four percent of participants tested partners using the INSTI kit (mean 1.6 times; SD 2.2 times).  Participants liked the tests because they were easy to use, gave quick results, and allowed dual HIV/syphilis testing. Participants stated that the blood-based nature of the test was a slight barrier. |
| **Baraitser 2019** | Descriptive quantitative (service evaluation)  N=1466  **PrEP-C.C.:** HIV testing (ordering tests, user to provider results sharing, HIV risk assessment)  **Modality:** Website  **Location:** England and Wales (UK) | The study evaluated HIV self-testing and self-sampling through an online sexual health service (SH24). Participants were directed to the study website where they completed a baseline risk assessment and chose the type of test they wanted to receive which were then sent to them. Participants were able to report their results via the study’s website. | HIVSTs provided = 984 (67.1% of total tests provided), known results = 563 (57.2% of HIVST provided).  HIV self-sample kits provided = 482 (32.9% of total tests provided), known results = 260 (53.9% of self-sample kits provided).  No new HIV diagnoses were observed. |
| **Bauermeister 2015** | Randomised controlled trial  N=130  **PrEP-C.C.:** HIV risk assessment  **Modality:** Website  **Location:** USA | Evaluation of the feasibility, acceptability and efficacy of an online intervention that allows young GBMSM to identify HIV testing locations filtered based by their specific structural needs. The website offers an online HIV risk assessment as part of this process. | 31% of participants reported booking an appointment to test for STIBBVs (no significant difference between intervention (32.4%) and control (27.8%)).  32.4% of the intervention group (all who booked a test) were tested for STIBBVs compared to 22.2% of the control group – not statistically significant but the authors report this as a clinical significance. |
| **Biello 2021a** | Focus group and descriptive quantitative (pilot)  N=28 qual; 11 feasibility  **PrEP-C.C.:** HIV testing (ordering tests); PrEP education  **Modality:** Smartphone app  **Location:** USA | Description of the development and evaluation of MyChoices – a smartphone app that aimed to increase HIV testing and PrEP use among young GBMSM. MyChoices was informed by social cognitive theory and provided a way of ordering self-tests as well as information about HIV and PrEP. The PrEP education in this instance was mainly considered to be offline apart from the ability to locate PrEP providers within the app. | **Qual:** Participants felt there needed to be an obvious usefulness in order to engage with the app. Participants liked that the app was not obviously about HIV or PrEP. Language that was considered to be “too academic” was viewed as a barrier to use.  **Feasibility study:** Participants used the app on average 8 times. 10/11 used the app to order a HIV or STI test and safer sex supplies; 7/11 used the app to locate HIV/STI testing centres or PrEP providers. Mean SUS score was 71 (SD 11.8).  82% said it was useful; 73% were very satisfied; 91% would recommend it; 82% would be likely to use it again if it became publicly available. |
| **Biello 2021b** | Descriptive data from a randomised controlled trial – categorised as descriptive quantitative  N=80  **PrEP-C.C.:** HIV testing (ordering tests; provider to user results)  **Modality:** Smartphone app  **Location:** USA | The study provided HIVST and STI self-sample kits via the LYNX and MyChoices apps. The apps allowed users to order self-test or self-sampling kits for HIV, syphilis, gonorrhoea and chlamydia. Results are sent to participants via email or the app if negative and over the phone if positive. | 76% of the participants who completed the follow-up survey had been tested for HIV in the study period. 54% reported using at least one HIVST in the study period. Average time for results to be delivered after sample collection was 10 days.  93% reported that it was 'not at all' or 'a little' difficult to order HIV/STI self-tests.  87% of app users reported that it was 'extremely' or 'very' helpful to order the kits and other safer sex supplies through the app. 80% reported that it would be very convenient to use HIV self-tests in the future and that they felt extremely confident in using the test correctly. |
| **Chan 2021** | Descriptive quantitative  N=350  **PrEP-C.C.:** HIV testing (ordering tests; pre- and post-test counselling)  **Modality:** Website, social media, video call  **Location:** China | Evaluation of an online HIV self-test service where participants received a free self-test and booked an appointment via a phone call or social media.  The appointment was conducted over video call with staff trained in motivational interviewing who delivered pre-test counselling, real-time instructions on how to administer the self-test, and post-test counselling and linkage to care. | 40.4% of new users and 63.1% of existing users ordered an HIVST during the study period. 58.6% of new users and 79.4% of existing users tested for HIV during the study period.  Four people were newly diagnosed with HIV.  123 completed the follow-up. Satisfaction with the different procedures of the online HIVST among this group ranged from 88.8% to 96.8%, and 72.0% to 97.6% of participants felt that the counselling component was helpful in understanding different aspects of HIV testing (e.g. window periods). |
| **Chasco 2021** | Non-randomised quantitative and semi-structured interviews  N=77  **PrEP-C.C.:** HIV testing (ordering tests); renal monitoring (order tests)  **Modality:** Video call, email  **Location:** USA | The study evaluated the addition of self-sampling to a telehealth program for PrEP (IowaPrEP). During a video-consultation with their pharmacist, or via email, participants could request a self-sampling kit for HIV, STIs and renal function (creatinine) to be posted to them or arrange for the participant to attend a clinic to have these tests completed. For those opting for the self-samples, the samples would be posted to the lab for analysis. | **Quant:** Participants who ordered self-sampling kits had 207 lab monitoring episodes during the study period, 79 of which were self-sampled. HIV samples were completed at 100% of the clinic visits and 83.5% of the self-samples. Adequate sample for creatinine analysis was obtained in 91.9% of the clinic visits and 81% of the self-samples.  **Qual:** Factors affecting choice: self-efficacy; time/convenience; privacy/confidentiality; method of delivery; and cost consideration. |
| **Cohen 2017** | Non-randomized quantitative (retrospective cohort)  N=1460  **PrEP-C.C.:** HIV testing (provider to user results)  **Modality:** Website  **Location:** USA | Evaluation of the effectiveness of Healthvana, an online portal for accessing STI test results including HIV. | Mean days between test and treatment for those accessing Healthvana was 10.68 compared to 12.55 for non-Healthvana participants. The mean days between test and notification of results was 6.86 for those accessing Healthvana compared to 8.77 for those not accessing Healthvana. There was no significant difference in days from notification to treatment. |
| **Daniels 2016** | Descriptive quantitative (cross sectional survey)  N=37  **PrEP-C.C.:** HIV testing (user to provider results)  **Modality:** Email  **Location:** USA | The study evaluated the acceptability of an intervention where people would complete an HIV self-test, take a photo of the completed test and send this to the research team. | Preferred method of sharing results were: email (37.8%); text message (21.6%); in person (16.2%); taking a picture and sending it (13.5%); in writing (5.4%); mailing the swab (2.7%); and by phone (2.7%).  Level of comfort taking a picture of the HIVST then texting the researcher: very comfortable (56.8%); somewhat comfortable (13.5%); neutral (10.8%); somewhat uncomfortable (5.4%); and very uncomfortable (13.5%). |
| **De Boni 2019** | Descriptive quantitative (feasibility)  N=4800  **PrEP-C.C.:** HIV testing (ordering tests)  **Modality:** Website  **Location:** Brazil | The study described the development and feasibility of a free, anonymous, internet-based HIVST strategy. The website included information about HIV, the option to order an HIVST, and an HIV risk assessment tool. Participants who met the study’s criteria could then arrange to pick up a kit from a pharmacy. Participants had the option of returning results via the study website or by mail. | Of the 2526 HIVSTs ordered, 21.5% reported their results. Of these, 23 were reactive and 11 were invalid.  Against the study’s set feasibility success outcomes:  Participants who received an HIVST after starting the online process = 65% (success indicator of ≥60%). Individuals who collected their HIVST from the pharmacy within two weeks of ordering = 38.5% (success indicator of ≥50%). HIVSTs distributed during the first 12 months = 2526 (success indicator of ≥1000). Proportion of HIVST results returned = 21.4% (success indicator ≥20%). Proportion of positive tests followed-up with confirmatory testing = 88.2% (success indicator ≥50%). |
| **Elliot 2016** | Descriptive quantitative (service evaluation)  N=17,361  **PrEP-C.C.:** HIV testing (test ordering; user to provider results sharing; HIV risk assessment), PrEP eligibility assessment (provisional)  **Modality:** Website  **Location:** England | The study evaluated Dean Street @ Home – a service that offers free at-home HIV self-sample kits, online HIV risk assessment, and PrEP recommendation. | 36% of those who completed the assessment had never been tested for HIV previously. 45% of those who completed the assessment were deemed at increased risk of HIV acquisition.  93% of people who clicked for more info actually requested a test. 55% returned their samples. 122 samples (121 people) were reactive, 82% were confirmed (14 false reactive, 14 already diagnosed; 11 unconfirmed). Linkage to care was 88%. 23% of people diagnosed with HIV through DS@H had a CD4 count <350, compared to 32.5% of traditional care. |
| **Finkenflügel 2019** | Descriptive data from a randomised controlled trial – categorised as descriptive quantitative  N=374  **PrEP-C.C.:** PrEP adherence monitoring  **Modality:** Smartphone app  **Location:** The Netherlands | The study explored participants’ use of the AMPrEP app. The app asked participants questions daily about adherence and sexual behaviour, and provided a visualisation of self-reported pill taking and sexual activity. | The percentage of participants who reported data 27 to 30 days per study month decreased over time (p<0.001).  The median number of pills taken according to the app tended to be lower than reported in the questionnaire for those on an event-based regimen – this was significant at month 9 and the questionnaire was administered every three months. |
| **Fuchs 2018** | Non-randomised quantitative and focus groups  N=56  **PrEP-C.C.:** PrEP adherence support  **Modality:** Email  **Location:** USA | The study evaluated the feasibility, acceptability and adherence effects of weekly bi-directional SMS- or email-based adherence support for PrEP. | 32.1% opted for email. 63% preferred messages in the morning, 30% in the afternoon. 80% preferred their message at the start of the week (Sun or Mon).  3 instances of "not okay", 1 requested clinical consultation due to side effects; 2 just wanted to see what would happen.  14% of the email users did not respond compared to 9% of the SMS users (p<.009). Response time for SMS was mean 4.4hrs (range 0.01-46.7) v email mean 6.1hrs (range 0.02-43.8) (p=.03). 56% said the messages were helpful. |
| **Gilbert 2017** | Descriptive quantitative (pilot)  N=868  **PrEP-C.C.:** HIV testing (online risk assessment, provider to user result sharing)  **Modality:** Website  **Location:** Canada | The study described the use, testing outcomes, and user characteristics of GetCheckedOnline. GetCheckedOnline allows participants to complete a risk assessment which signals which tests they are advised to complete. The participant takes this form to the GCO lab site. Negative results can be accessed on the website. | 695 assessments were completed by 506 clients.  10 (3.1%) had a positive STI diagnosis (not reported what STI(s)). 12.5% were first testers. |
| **Gilbert 2018** | Descriptive quantitative (cross-sectional survey)  N=394  **PrEP-C.C.:** HIV testing (online risk assessment, provider to user result sharing)  **Modality:** Website  **Location:** Canada | The study compared users’ experiences of testing barriers between in person clinic attendees and GetCheckedOnline clients. GetCheckedOnline allows participants to complete a risk assessment which signals which tests they are advised to complete. The participant takes this form to the GCO lab site. Negative results can be accessed on the website. | When compared to clinic attendees, GetCheckedOnline clients were more likely to be older (median age: clinic = 30; GCO = 35); test more frequently (67% GCO tested regularly compared to 39% of clinic attendees); delay testing due to clinic distance (clinic = 9%; GCO = 28%); delay due to waiting times (clinic = 54%; GCO = 76%); report embarrassment with testing (GCO = 16%; clinic = 6%) and discussing sexual history (GCO = 19%; clinic = 5%); fear judgement from healthcare workers (GCO = 30%; clinic = 15%). GCO clients were less likely to report that clinic times were convenient (GCO = 77%; clinic = 59%); and more likely to report experiencing long wait times in clinic (GCO = 50%; clinic = 17%). |
| **Gilbert 2019a** | Critical appraisal/review  N=46 websites  **PrEP-C.C.:** PrEP education  **Modality:** Websites  **Location:** Canada (limited to Canadian websites) | The study critically evaluated information included on Canadian HIV websites relating to HIV risk factors and prevention strategies. The evaluated how this information was presented, and its readability, usability and interactivity. | 76% were run by community based organisations. 3 had an online risk assessment tool. Flesch-Kincaid median [IQR] score: 62.5 [52.0-58.4]. LIDA Usability mean(SD) [possible points]: clarity of design 14.1 (2.1) [18]; consistency of design 8(1) [9]; functionality 10.7(1.7) [15]; engagement 3.8(1.2) [12]; Total 36.6(4.3) [54]. 38% used colloquial language, 88% used plain, and 40% used technical (does not add up to 100% as some sites used more than one style). 40% focused on HIV only, 38% on sexual health including HIV; 20% on general health including HIV; 2% was other. PrEP was mentioned in 10% of sites. |
| **Gilbert 2019b** | Non-randomised quantitative  N=19,497  **PrEP-C.C.:** HIV testing (online risk assessment, provider to user results)  **Modality:** Website  **Location:** Canada | The study compared rates of repeat testing among people using GetCheckedOnline with people who used STI clinics. GetCheckedOnline allows participants to complete a risk assessment which signals which tests they are advised to complete. The participant takes this form to the GCO lab site. Negative results can be accessed on the website. | For GCO, there were 1951 testing episodes across 1093 clients. For the STI clinics, there were 39,357 testing episodes across 18,404 clients. STI clinic clients had repeat tested 1.53 times/person year and GCO clients had repeat tested 1.87 times/person year (this was statistically significant).  Most GCO clients only tested through GCO. Of the 272 who tested at both GCO and an STI clinic, 87% were tested at a clinic first, 13% were tested through GCO first. GCO clients were more likely to be White, female, men who have sex with women, have had a previous STI, and a partner living with HIV. GCO clients were less likely to be GBMSM. |
| **Guinness 2018** | Descriptive quantitative (pilot)  N=126  **PrEP-C.C.:** PrEP education  **Modality:** Email  **Location:** USA | The study described an intervention where a one-time secure email or letter providing information about PrEP and linkage to PrEP care was sent to clinic attendees. | Median time from STI to outreach was 164 days (IQR: 98-222).  78.8% of those who were sent the message read it. No differences in reading of message by age, gender or race/ethnicity.  12.4% of those emailed were linked to PrEP care; 11(91.7%) of those filled a prescription. All those who were linked to PrEP were from the email group. |
| **He 2018** | Descriptive quantitative (proof of concept)  N=500  **PrEP-C.C.:** HIV testing (provider to user results sharing)  **Modality:** Website  **Location:** China | The study evaluated the feasibility of providing anonymous urine-testing for HIV at drug stores. After the sample has been mailed or handed into a pharmacy and processed, participants can enter a unique code that came with the kit on the study website and access their results. | 430 (63%) of the kits were submitted for analysis. 70 (16.3%) of the tests were reactive. 94.3% of the people with a reactive result and 83.7% of the people with a negative result accessed their results online.46 of the 70 people (65.7%) with reactive tests were followed up, 40 agreed to have their results confirmed, 39 of these were confirmed to be living with HIV, 37 of these were new diagnoses, 28 were given free ART. |
| **He 2019** | Descriptive quantitative  N=957  **PrEP-C.C.:** HIV testing (provider to user results sharing)  **Modality:** Website  **Location:** China | The study described a service where participants were offered anonymous HIV testing via urine samples which are distributed via vending machines on university campuses. After the sample has been mailed or handed into a pharmacy, participants can enter a unique code on the study website and access their results. | 957 tests were distributed, 378 were returned (376 of which were valid for analysis). 255 participants accessed their results. 65.9% of participants whose test was negative accessed their results. 100% of participants whose test was reactive accessed their results (n=7). 88.9% of the kits were dispensed between 9pm and midnight.72.2% of the kits were dispensed in dorms, 27.8% in teaching buildings. |
| **Hoth 2019** | Descriptive quantitative  N=186  **PrEP-C.C.:** HIV testing (risk assessment; appointment booking); renal monitoring (appointment booking); PrEP education; PrEP adherence (counselling); PrEP eligibility assessment  **Modality:** Video call  **Location:** USA | The study described Iowa TelePrEP, a service where people who would benefit from PrEP are referred to a pharmacist who conducts a video consultation with the client. The video consultation involves a PrEP eligibility assessment (including HIV risk assessment), PrEP education, and PrEP adherence counselling. If the client meets the required criteria, the pharmacist will book them into a clinic of their choosing for tests (STIBBV and renal function), and will send their PrEP to the client by post. | There were 186 referrals which resulted in 127 initial video-consultations (all completed within 40 days of the referral; 78% of these were completed within two weeks of referral; and 53% within one week).  Retention was 60% (50/83) at 180 days (for clients who had enough time for follow-up to be measured).  Completion of blood tests was 96%.  1 client was newly diagnosed with HIV at baseline.  Of the 167 eligible creatinine visits, 98% (n=164) were complete. |
| **Hottes 2012** | Focus groups  N=39  **PrEP-C.C.:** HIV testing (HIV risk assessment; provider to user results)  **Modality:** website  **Location:** Canada | The study explored the acceptability of internet-based STI/HIV testing where users complete an HIV/STI risk assessment online which suggests which tests the user should get at the study’s lab sites. This list of tests can be printed out. Test results are uploaded to the study website and can be accessed by the user. | Participants were concerned about giving information online and the possibility of people entering others’ details online maliciously (could be remedied with an email verification). Participants only wanted to provide essential information where each question explained by the information was necessary.  Participants were concerned about positive tests and how to interpret the tests. Participants preferred that positive HIV test results should be communicated over the phone.  Participants wanted a similar level of service as face-to-face and that online care may not be suitable for first time testers. |
| **Huang 2016** | Descriptive quantitative  N=122  **PrEP-C.C.:** HIV testing (test ordering)  **Modality:** Website  **Location:** USA | The study evaluated an HIV testing program that linked social media advertisements with an online test requrest system. | 81 (66.4%) confirmed to have received a self-test. No vending machine codes were actually used. 93% received their test by mail and 7% by voucher. 96.5% found the self-test easy or very easy, 68.4% would prefer self-testing among other testing choices in the future; 96.5% reported testing negative; 3.5% reported newly testing positive for HIV but reported seeking care. |
| **Hughes 2021** | Qualitative interviews  N=31  **PrEP-C.C.:** HIV testing (risk assessment; provider to user results sharing); PrEP eligibility assessment; PrEP education; PrEP adherence counselling; PrEP prescription (notification; user provides mailing information)  **Modality:** Website  **Location:** USA | The study described Nurx, a website that offers internet-based PrEP. When clients create a profile on the website, they are asked to complete an HIV risk assessment. This is reviewed by Nurx staff and potentially a clinician. Electronic orders are sent to a lab where the client presents for testing. This data is relayed to Nurx who review the results and notify the client via the Nurx website. The patients then receive further PrEP education and adherence counselling via the website and a 3-month supply of PrEP is sent to them. | Most participants were extremely satisfied with the service and reported having to “overcome scepticism” as the service appeared too good to be true or a departure from the care they were used to receiving. Participants found the web-based nature of the service convenient and felt it was able to strike a balance between efficiency (simplicity, speed, convenience) and “humanity” (personalised, responsive, feeling of a connection/care). |
| **Jackman 2018** | Focus groups and descriptive quantitative  N=35 (FGs); N=380 (survey)  **PrEP-C.C.:** HIV testing (provider to user result sharing)  **Modality:** Website  **Location:** USA | The study explored how the introduction of electronic patient health records may impact sexual health-related communication between sexual partners (i.e. sharing evidence of testing history). | **Qual:** The type of conversations that the electronic records may help facilitate usually take place at the start of a relationship. The records could make conversations about STI statuses easier.  **Quant:** 60% said ePHR will help initiate conversations, 225/354 felt ePHR would improve communication; 235/254 believed ePHRs would increase confidence in STI testing info shared by partner; 197/354 said it would make discussing risk with new partners and 195/354 with existing partners.  85/354 said it'd be awkward soliciting partners' records. 154/354 said it'd be difficult sharing results with partner. 138/354 preferred disclosing a positive result without the ePHR, while 81/354 preferred with. 268/354 believed that potential partners would refuse to share. |
| **Jin 2018** | Descriptive quantitative (feasibility)  N=879  **PrEP-C.C.:** HIV testing (HIV risk assessment; test ordering; user to provider results sharing)  **Modality:** Website  **Location:** China | The study assessed the feasibility of the Easy Test model – an internet-based HIVST service where users completed an online HIV risk assessment and ordered an HIV self-test kit. Users were then able to report their test result online and be linked to care if needed. | 40% reported never having been tested. 77.7% submitted a photo of completed test. 14.3% of these were positive. 72.4% of those who were diagnosed were linked to treatment within a month. 42.9% of those who submitted photos were first time testers. Within the first time testers, 18.8% HIV prevalence. HIV diagnosis was related to receptive anal intercourse and vers, greater numbers of sexual partners, inconsistent condom use, no condom in most recent AI, no knowledge of sexual partner's status, and never been married. |
| **Kecojevic 2018** | Content evaluation  N=217  **PrEP-C.C.:** PrEP education  **Modality:** YouTube videos  **Location:** N/A | The study describes a content evaluation of YouTube videos that provide information about PrEP. | Collective views = 2369003 (single CDC had over 1.2 million views alone - an animation w/voiceover 07.01.2016, 2mins 51 secs). 30.4% (n 66) were from institutions and 29% (n 63) from consumers; 22.1% (n 48) from CBOs, and 18.4% (40) from media. No significant difference in length between source but sig diff in views (p .003) and comments (p<.001): institutions had fewer views than consumer and media; consumer vids had more comments than institutions and CBOs, and media had more than institutions. Overall: 82.9% (n=180) defined PrEP; 49.3% (n=107) described how PrEP works; 60.8% (n=132) described who could use PrEP; 23% (n=50) promoted PrEP as a safe option; 32.3% (n=70) discussed side effects; 35.9% (n=78) described how to obtain PrEP; 27.6% (n=60) discussed the cost of PrEP; 83.4% (n=181) promoted the use of PrEP. |
| **Knight 2019** | Semi-structured interviews  N=37  **PrEP-C.C.:** HIV testing (online risk assessment, provider to user results)  **Modality:** Website  **Location:** Canada | The study explored GBMSM’s experiences with, and preferences for, GetCheckedOnline and clinic-based STI/HIV testing. GetCheckedOnline allows participants to complete a risk assessment which signals which tests they are advised to complete. The participant takes this form to the GCO lab site. Negative results can be accessed on the website. | Participants preferred GCO over clinic-based testing as it was more convenient and quicker. Participants said that GCO gave them autonomy, control and they could act immediately; it enhanced privacy and reduced thoughts of being judged.  Many said that they were aware of their sexual risks and viewed counselling as unnecessary. GCO changed frequency of testing due to the email reminders. Participants found the emails also improved understanding around risk. Participants felt that being able to access results online reduced anxiety and uncertainty around results. GCO was seen as a complement to clinic based care - that participants used GCO for STI/HIV testing but clinic for other care. Some participants preferred clinic-based testing as there was a larger variety of services not available through GCO. Some preferred clinicians doing the sampling procedures. Some felt that GCO was redundant given that they already go to their doctor to receive other care. Some preferred sexual health clinics over the lab environment of GCO. Some participants worried about missed opportunities via GCO. |
| **Koester 2020** | Qualitative interviews  N=31  **PrEP-C.C.:** HIV testing (risk assessment; provider to user results); PrEP eligibility assessment; PrEP education; PrEP adherence counselling; PrEP prescription (notification; patient provides mailing information)  **Modality:** Website  **Location:** USA | The study explored the acceptability of lab monitoring for current and potential future user of Nurx. See Hughes 2021 for a detailed overview of Nurx. | Participants found presenting to a clinic to testing or to drop of specimens acceptable. The authors note that Nurx has a variety of sites with long opening hours. Participants felt that the need to test every three months helped to create a healthy sexual health screening structure within their lives. |
| **Lee 2020** | Content analysis  N=559 questions  **PrEP-C.C.:** HIV testing (counselling); PrEP education  **Modality:** Website  **Location:** South Korea | In the context of an online HIV counselling website, the authors identified questions frequently asked by users relating to HIV and PrEP. | The categories questions identified were as follows: HIV testing, self-perceived HIV risk and sexual behaviours, positive and negative emotional state, and treatment and prevention. |
| **Lessard 2019** | Focus groups  N=21  **PrEP-C.C.:** HIV testing (provider to user and provider to users’ partner(s) results notification)  **Modality:** Smartphone app  **Location:** France | The study described the acceptability of the WeFLASH© partner notification app. The app allows people to ‘flash’ each other which connects their apps. If a person is diagnosed with HIV or an STI, their lab results are added to the app and this notifies any partners that they should seek testing. | Potential benefits of the service are that it could allow better patient notification, customized linkage to care, and transferrable data.  Potential risks could be privacy and confidentiality, changes in sexual behaviour, and fairness. |
| **Liu 2021** | Focus groups, descriptive quantitative, and semi-structured interviews  N=54 (focus groups); N=20 (pilot); unclear if there is overlap.  **PrEP-C.C.:** PrEP adherence monitoring and support  **Modality:** Smartphone app  **Location:** USA | DOT monitors and supports PrEP adherence. The app captures data through the phone’s camera (confirmed by user via app). Participants receive daily reminders to take their PrEP and receive a prompt if the dose is late or missed. Data is uploaded in real-time to a secure cloud that can be accessed by study staff. The app allows users to track their sexual behaviour which is integrated with the PrEP adherence data to feedback to the participant if/when they are/aren’t protected. | Participants praised the ability to track their level of protection. Median SUS score was 85/100 at week 4 and 80/100 at week 8. 84% reported that they were likely to use the app again in the future.  Among 1198 expected doses, 80% were confirmed, 6% were self-reported, and 14% were missed. Median PrEP adherence was 91%. |
| **MacGowan 2019** | Randomised controlled trial  N=2665  **PrEP-C.C.:** HIV testing (ordering tests, user to provider result sharing)  **Modality:** Website  **Location:** USA | Participants were randomised to receive the intervention or the control (routine care). Both groups completed the baseline survey and the HIVST kits were mailed to the intervention group. Quarterly surveys were completed by all and the intervention group could order additional kits at this point. Participants reported their results via the study website. | A higher proportion of participants in the intervention group reported testing 3 or more times during the trial than in the control group (76.6% vs 22.0%; p<0.01).  The largest difference in HIV diagnoses was during the first three months (0.9% intervention vs 0.1% control; p<0.01). |
| **Maksut 2016** | Descriptive quantitative (test of concept)  N=20  **PrEP-C.C.:** HIV testing (counselling)  **Modality:** Video call  **Location:** USA | The study described a service that provided video-call-based HIV self-test support (pre-test counselling, guidance/support during test, post-test counselling). Eligibility was assessed via phone call and their test kit was sent. When completing the test, participants attended a video call with staff who provided counselling. | All participants found the service satisfactory and would access in it again if it were made available. They would also recommend it to a friend. 72% of participants would prefer their next test to be conducted in this way. All tests were negative. |
| **Manavi 2017** | Descriptive quantitative  N=5301  **PrEP-C.C.:** HIV testing (HIV risk assessment; HIV testing kit ordering)  **Modality:** Website  **Location:** England (UK) | The study described the Umbrella Health website which contains an assessment that determines what type of test kit(s) is needed. Patients then have this delivered/pick it up, complete it and post back to the clinic. Results provided via SMS. | Women were more likely to return kits than men (61.2% v 53.1%, p<.001). Only 1 trans patients returned their kits (n=10 total). GBMSM (62.5%) and women had a similar return rate (61.3% for non-anorectal, 59.6% for anorectal included). Heterosexual men less likely at 49.6% (p<.001).  People preferred to have kits sent to their home 77.49% (rather than 11.92% at clinic and 10.58% at pharmacy). |
| **Menza 2021** | Descriptive quantitative  N=233 participant who ordered a total of 248 kits  **PrEP-C.C.:** HIV testing (test ordering)  **Modality:** Website  **Location:** USA | The study described Oregon’s state-wide pilot HIVST program. Participants were able to order HIVST kits from the study website which were delivered by mail. | 22% of the participants lived in rural zip codes.  73% of the 150 kits assigned to the study to begin with were ordered within the first 24 hours of implementation.  Participants said that the service was convenient and enhanced privacy. |
| **Mitchell 2018** | Descriptive quantitative (single arm trial)  N=12  **PrEP-C.C.:** PrEP adherence monitoring  **Modality:** Smartphone app  **Location:** USA | The study examined the feasibility and acceptability of mSMART for PrEP – real-time camera-based event-monitoring. | Adherence increased for 30% of participants and stayed the same for the remaining 70% stayed the same. Those who didn't change, their PrEP adherence was already considered efficacious at baseline. Participants logged a dose 91% of days. 88% used the camera function. 40% did not miss any days.  On a scale of 1 (not at all) to 4 (extremely) (mean (SD)): satisfaction (2.80 (.63)); usability on a daily basis (3.50 (.53)); willingness to recommend (2.70 (.82)); user-friendliness (2.80 (.79)); difficultly to learn how to use (1.20 (.42)). SUS mean 68.25 (15.10); having a SUS score of 68 was deemed to indicate acceptability, 60% reached this.  Participants said that the app would be most useful for people initiating PrEP and those with adherence issues. |
| **Page 2019** | Non-randomised quantitative  N=550  **PrEP-C.C.:** HIV testing (order test; provider to user result sharing)  **Modality:** Website  **Location:** England | The study compared the feasibility of dry blood spot and micro tube HIV self-sample kits where participants ordered the kits online, sent them into a lab, and received their results online. | 68.7% of MTs were returned compared to 66.5% of the DBS.  98.8% of DBS were successfully processed compared to 55.7% of the MTs (p<0.001). |
| **Phanuphack 2018** | Non-randomised quantitative (self-selecting trial)  N=571  **PrEP-C.C.:** HIV testing (counselling, test ordering, appointment booking, user to provider and provider to user results)  **Modality:** Website, video call  **Location:** Thailand | The study explored the acceptability and feasibility of an online HIV testing and linkage service (myhealth.adamslove.org (electronic health records)) for HIV testing: offline (face-to-face care and referred to the website for info post-care); online (supervised HIV self-testing entirely remotely with virtual care); and hybrid (online pre-test counselling and appointment booking, HIV test completed at clinic). | Attitudes towards HIV testing: 13.7% afraid of needles; 36.1% concerned about confidentiality; 14.7% less benefit than harm of knowing HIV result; 54.5% wanted a rapid result; 32.5% wanted home testing, 56.6% thought HIV testing was good way of looking after one's health.  Online pre-test counselling was associated with having monthly income between 429 and 857 USD (OR 2.31, p=.04), having had two or more HIV tests in the past (OR=2.57, p=.04), and among MSM identifying inconvenient service hours as a barrier to testing (OR=2.92, p=.008).  Being trans (OR=6.66, p<.001), spending 4-8 (OR 2.82, p=.002) or >8 hours on social media (OR 2.33, p=.04), having preference for online services (OR 5.73, p<.001) and a preference for home-based HIV testing (OR 6, p<.001) increased patient's likelihood of choosing online HIV testing and post-test counselling.  Having a preference for immediate confirmatory HIV testing and ART (OR .32, p=.003), wanting a rapid test (OR .3, p<.001), having concerns around quality (OR .42, P=.01) or clinical hygiene (OR .31, p<001) reduced likelihood of access online testing and post-test counselling. |
| **Polilli 2016** | Descriptive quantitative  N= “about 6000” - exact numbers not reported  **PrEP-C.C.:** HIV testing (HIV risk assessment; appointment booking)  **Modality:** Website  **Location:** Italy | The study evaluated the efficacy of a web-based HIV testing initiative where participants could complete an HIV risk assessment then book an appointment for testing at one of six clinics. | 28 (0.92%) of the people tested were diagnosed with HIV; 26 (92.8%) were linked to care. The service was seen to be cost effective given that the number of diagnoses exceeded the 1/500 threshold. |
| **Refugio 2019** | Non-randomised quantitative (feasibility)  N=25  **PrEP-C.C.:** PrEP education; PrEP eligibility assessment; PrEP adherence  **Modality:** Website  **Location:** USA | Investigated the feasibility of PrEPTECH. Participants could register on the study website and complete a baseline PrEP knowledge quiz. Home testing for chlamydia and gonorrhoea was delivered to the participant’s home but they had to attend a lab to test for HIV testing. When the safety criteria were met, a 90-days’ supply of PrEP was delivered to the participant’s home. At 30, 90 and 180 days, physicians completed a phone consultation with the participants and issued further PrEP. At 90 and 180 days, participants were invited to complete a follow-up survey in which they reported their adherence. | The number of participants who felt uncomfortable going to a clinic for PrEP: 66.7% at 90 days, 72.2% at 180 days (no n reported). 85% agreed that PrEPTECH was better way for gay/bisexual men to get PrEP. 88% reported that PrEPTECH was very easy or extremely easy to use.  All participants reported that PrEPTECH was very or extremely fast and convenient compared to other routes of PREP access.  Half reported hearing of others experiencing stigma from taking PrEP. However, only 14.3% at day 90 and 10.5% at day 180 had experienced PrEP stigma personally - mostly from the gay community but healthcare providers, family, friends, and strangers were also cited as sources. Median days from online eligibility to first shipment of PrEP was 46 days (range = 21-133 days). |
| **Rosengren 2016** | Descriptive quantitative (cross-sectional survey)  N=125  **PrEP-C.C.:** HIV testing (test ordering)  **Modality:** Website  **Location:** USA | The study described how they distributed HIV self-test kits by advertising through Grindr. Participants were linked to the study site where they could order an HIVST. | 33 (59%) said HIVST kit was very easy to use, 34% said it was easy; 4% neutral; 4% hard; 0% very hard.  54 (96%) had a negative HIVST result.  When comparing the HIVST kit and clinic testing, 35 (63%) preferred HIVST kit; 8 (14%) somewhat preferred the HIVST kit; 6 (11%) neutral; 3 (5%) somewhat preferred the clinic; and 4 (7%) preferred the clinic. |
| **Salway 2019** | Non-randomised quantitative  N=352  **PrEP-C.C.:** HIV testing (online risk assessment, provider to user results)  **Modality:** Website  **Location:** Canada | The study compared GetCheckedOnline clients’ HIV test knowledge and risky sexual behaviour with clients accessing in clinic testing. GetCheckedOnline allows participants to complete a risk assessment which signals which tests they are advised to complete. The participant takes this form to the GCO lab site. Negative results can be accessed on the website. | GCO users demonstrated higher HIV post-test knowledge at baseline, this became non-significant in adjusted analyses.  GBMSM, clients with university degrees, clients who had lived in Canada for more than 10 years, and English speakers had higher HIV post-test knowledge than their equivalent counterparts (p<0.05). |
| **Siegler 2019** | Descriptive quantitative  N=58  **PrEP-C.C.:** PrEP eligibility assessment; HIV testing (instructional video)  **Modality:** Website  **Location:** USA | The study described PrEP@HOME. Participants received a self-sampling kit at their home. Once this was returned, processed and an online behavioural surveillance survey was complete, another PrEP prescription was issued (if the clinician deemed this safe and appropriate).  While the service provides comprehensive PrEP care, it appeared that only the behavioural surveillance and instructional video were online. | 14 (25.45%) had mildly low GFR. From the 57 patients, 4 required standard care. 53 (93%) were able to have their prescriptions renewed via PrEP@HOME only. Participants rated PrEP@HOME kit as good on the SUS scale (mean score (SD): 76.91 (18.4)). 85% indicated they would opt for PrEP@HOME over in clinic visit within the following year. |
| **Stekler 2018** | Non-randomised quantitative  N=48  **PrEP-C.C.:** PrEP education  **Modality:** Video call  **Location:** USA | The study described the use of video call within PrEP initiation appointments. Participants had the option of having the physician present in person or via video call. Regardless, an HIV counsellor was present to collect the necessary specimens. | There was no statistically significant difference in the proportion of participants prescribed PrEP in the video call arm (70%) compared to when the physician was present (79%). 40% of the participants in the video call arm attended a three-month follow-up appointment compared to 87% of the standard care patients (p=0.05). |
| **Stephenson 2020** | Randomised controlled trial  N=202  **PrEP-C.C.:** HIV testing (counselling and support, user to provider results)  **Modality:** Video call; website  **Location:** USA | The study evaluated the pilot study of Project Moxie – a service where participants attend a video call appointment with an HIV counsellor who provides real-time instructions and pre- and post-test counselling for people while they complete an HIV self-test. Participants in the control group (who received HIVST but no video call appointment) could report their HIVST results via the study website. | 100% of the control group ordered an HIV self-test, 91% reported their HIV test results via the study portal.  48% of those randomised into the intervention group opted to receive the intervention - all of whom ordered a test and received video-chat counselling. The self-test was performed in the session in 96% of cases. All had a non-reactive result.  Satisfaction was high (overall satisfaction = 98.3%). |
| **Sullivan 2017** | Descriptive quantitative  N=121  **PrEP-C.C.:** HIV testing (HIV risk assessment; test ordering), PrEP eligibility assessment.  **Modality:** Smartphone app  **Location:** USA | The study assessed the usability and acceptability of HealthMindr – an app with HIV risk assessment, PrEP screening, PEP screening, HIV test comparison tool, and HIV test ordering. | App usage data was available for 109 (90%) of participants.  100% completed baseline assessment, just over 80% ordered a kit or condoms. 64% of kits ordered were OraQuick kits. 41% of all whom ordered a kit did so more than once. 4% of participants received reactive HIV test results. 8/86 eligible for PrEP and not already taking PrEP began taking PrEP. 6 mentioned that the app facilitated this.  66% felt that the app helped them stick to their HIV prevention plan. 71% said the app had a good mix of personal and professional language; 90% said the info was easy to understand; 86% were confident in the app security; 85% said the PIN was adequately safe; 84% said the app name and icon were not readily associated with HIV prevention.  Composite SUS score for usability was 73.4 (above average). |
| **Syred 2019** | Non-randomised quantitative  N=6253 users ordered 7550 tests prior to implementation; 7772 users ordered 9785 tests following implementation  **PrEP-C.C.:** HIV testing (test ordering)  **Modality:** Website  **Location:** England (UK) | The study described participants’ use of the ‘choose to test’ service. The service provided information on STIs and advice on regular testing. The service used an algorithm to recommend what tests people should order. People could then manually select and deselect tests as they wished. The tests were posted to them. | When compared to data for the same time period prior to implementing the ‘choose to test’ service, the positivity rate for gonorrhoea and chlamydia tests was constant. Too few HIV and syphilis tests were completed to analyse.  GBMSM were offered genital, rectal and oral chlamydia and gonorrhoea tests plus HIV and syphilis testing - in 17.2% of orders, people removed tests.  BME users were offered chlamydia, gonorrhoea and HIV testing, in 77.9% of order, people added tests. |
| **Wang 2018** | Randomised controlled trial  N=430  **PrEP-C.C.:** HIV testing (real-time counselling and support)  **Modality:** Website, video call  **Location:** China | The study evaluated the efficacy of an online intervention that provides real-time instructions and counselling for HIV self-testing in increasing HIV testing. After a baseline telephone survey, participants were randomised to be shown an online video about HIV or a video promoting the intervention. Participants who opted to use the intervention were sent an HIV self-test kit and made an appointment to receive the intervention via video call. | Those in the intervention group reported higher prevalence of HIV testing of any type at 6 months (89.8% v 50.7%; RR 1.77, NNT 2.56, p<.001).  Self-reported uptake of HIVST was 87.9% in the intervention arm compared to 2.3% in controls (p<.001).  81.3% felt that the content of the health promotion was clear. 71.4% believed the service was effective in reducing their risk behaviour. 78.5% expressed a desire to use the service again and 75% would recommend it to their peers in the next year. 47.8% would pay for HIVST-OIC at the cost of HK $100 per test in the next six months. |
| **Whiteley 2020** | Content review  N=61 websites; 58 YouTube videos  **PrEP-C.C.:** PrEP education  **Modality:** Websites, YouTube  **Location:** Global search (English language only) | The authors evaluated online content and YouTube videos about PrEP. Four criteria were used to appraise the websites: PrEP educational content, authority of PrEP website/video, cultural content, usability, and interactivity. | No website adequately addressed the four criteria used to appraise the sites. 3/61 websites had a perfect score for educational content and authority. 2/61 websites had a perfect score for cultural content. Very few websites had high interactivity scores (53/61 scored 25% or less) and 28 scored 50% or higher for usability. 13/58 YouTube videos had more than 10,000 views and 6 had more than 100 likes. |
| **Wilson 2017** | Randomised controlled trial (single blind)  N=2072  **PrEP-C.C.:** HIV testing (test ordering)  **Modality:** Website  **Location:** England | The study assessed the effect of SH24 (an online STI testing and results service (including HIV)) on uptake of STI testing, and STI cases diagnosed and treated. Participants were able to order their sampling kit online and their results were communicated via SMS. | 50% of intervention group and 26.6% of the control group had completed an STI test by week 6 (RR 1.87, p<.001) and 2.8% of the intervention and 1.4% of the control group had been diagnosed with an STI (RR 2.10, p=.079). Time to test was shorter in intervention group (28.8 days v 36.5 days, p<.001). Time to treatment was not significantly different (83.2 v 83.5 days p=.51). Median time from diagnosis to treatment was 2 days for the intervention and 4 days for control. 71% found the intervention acceptable. 88% of those in the intervention group tested via SH:24. |
| **Witzel 2019** | Randomised controlled trial and focus groups  N=1035 feasibility; 10 focus group  **PrEP-C.C.:** HIV testing (ordering tests)  **Modality:** Website  **Location:** England and Wales (UK) | The study assessed the feasibility an online HIVST randomised controlled trial’s recruitment strategy. Participants were randomised to receive a free HIV self-test, or not. The kits are ordered on the study website. | 76% of the people who registered were randomised. 98% of the participants who used the kit and completed the three-month survey found the instructions easy to understand; 97% found the test simple to use; and 97% reported a good overall experience.  Blood collection was a barrier to people who had never used an HIV self-test before. The intervention helped overcome geographic barriers and improved privacy. The intervention reduced anxiety for experienced testers (immediate results) and increased anxiety for others who were concerned about testing outside of a clinic setting (no support). |
| **Witzel 2021** | Randomised controlled trial and semi-structured interviews  N=118  **PrEP-C.C.:** HIV testing (ordering tests)  **Modality:** Website  **Location:** England and Wales (UK) | The study evaluated an online trial of HIV self-tests with a focus on trans participants. Participants were randomised to receive a free HIVST, or not. The kits are ordered on the study website. | In trans men, HIV testing uptake was significantly higher in the baseline test group (95%) than in the no baseline test group (29%) (p<0.001). Trans people in the repeat test group had a 3 times higher rate of repeat HIV testing compared to no repeat test group (p<0.0001). Acceptability was very high in the baseline test group. Perceived benefits included: increased autonomy, privacy, convenience and avoidance of discrimination and dysphoria often experienced in clinic. |
| **Woywodt 2014** | Descriptive quantitative (cross-sectional survey)  N=295  **PrEP-C.C.:** Renal monitoring (provider to patient results sharing)  **Modality:** Website  **Location:** England (UK) | The study evaluated Renal Patient View – a website where patients can view their renal test results. | Most users (78%) accessed the site 1-5 times/month. 81% used RPV to check creatinine levels. 92% said it was easy to use; 17% said they would have liked to have training. 2% did not think the information on the site was accurate. 32% were worried about accessing their results. 93% said it was a useful system. 80% said it helped them to monitor their kidney function. |
| **Wray 2018** | Randomised controlled trial  N=65  **PrEP-C.C.:** HIV testing (notification of test initiation)  **Modality:** Smartphone app  **Location:** USA | The study evaluated an intervention where “beacons” signalled when HIVST kits were opened. This prompted the research team to follow-up with the participant to see if further care or support was required. | Completion rate for monthly surveys was 89%. Attrition did not differ by arm, nor did survey completion rates. No reactive tests. 93.1% of participants reported using a study provided test. Sensors detected the kit opening 73.4% of the time.  All participants in the HIVST arms tested for HIV within the study period. |
| **Xia 2018** | Descriptive quantitative (feasibility)  N=3092 packs distributed; N=1977 packs returned  **PrEP-C.C.:** HIV testing (provider to user results sharing)  **Modality:** Website  **Location:** China | The study evaluated the feasibility of anonymous urine testing for HIV. After the sample has been mailed or handed into a pharmacy, participants entered a unique code that came with the kit on the study website and accessed their results. | 1911 samples were eligible for analysis, 145 were reactive, 104 were successfully followed-up, 10 of whom had previously been diagnosed with HIV. 1243 people accessed their results online. Of the 145 people with a reactive test, 112 (83%) accessed their results online. 94 (70%) of the people who had a reactive test were reached, 58 of whom sought confirmatory testing. |
