## Supplementary Material 5 for "Delivering HIV pre-exposure prophylaxis (PrEP) care online: A scoping review"

| **Author(s)** | **Acceptability** | | **Feasibility** | | |
| --- | --- | --- | --- | --- | --- |
|  | **Method of measuring acceptability** | **Level of acceptability** | **Uptake** | **Retention** | **Details on service delivery** |
| **Anand 2017a** | NR | NR | Reached 272,568 people in 3 months.  435 booked an appointment. | 325 (76.5%) checked in at one of the study sites. | The study identified 9 people who were unknowingly living with HIV.  The study reached people who were considered at high risk of acquiring HIV (22.8% had >5 sex partners in the last 6 months; 26.8% reported ever having an STI; 48.1% had sometimes or never used condoms in the last 6 months; and 30.2% were aware of all partners’ HIV status).  53.2% of the participants who were HIV-negative decided to initiate PrEP. |
| **Anand 2017b** | 110 (59.1%) participants completed a satisfaction survey which operated a 5-point Likert scale (higher number = higher satisfaction).  Mean (SD) reported. | Overall satisfaction = 4.4 (.68).  Design and interface = 4.34 (.78).  Consent and understanding = 4.58 (.57).  Ease of registration = 4.51 (.63).  Online data security = 4.64 (.53).  Ease of accessing lab results and post-test counselling summaries = 4.37 (.70).  Video chat quality for guidance = 4.71 (.47). | 489 people were assessed for HIV risk and introduced to the study arms.  186 (38%) enrolled in the study:  89 (47.9%) in the offline arm.  72 (38.7%) in the hybrid arm.  25 (13.4%) in the online arm. | Percentage of participants who revisited after baseline:  48.0% in the offline arm.  62.5% in the hybrid arm.  48.3% in the online arm. | The online arm seemed to be most effective at reaching those at higher risk of HIV given the percentage of participants in each arm newly diagnosed with HIV:  2.2% in the offline arm.  1.4% in the hybrid arm.  16.0% in the online arm.  The online arm also seemed to be very effective at reaching people who had never previously tested for HIV:  19.1% of the offline arm.  13.9% of the hybrid arm.  31.6% of the online arm. |
| **Balán 2020** | NA | NA | NR | NR | NR |
| **Balán 2021** | Cross-sectional survey and qualitatively via interviews. | On a 7-point Likert scale measuring the helpfulness of SMARTtest components (1 = not helpful; 7 = extremely helpful), mean scores varied from 5.74 (locating clinics using zip codes) to 6.46 (video instructions).  On a 5-point Likert scale measuring functionality (1=completely disagree; 5 = completely agree), mean scores varied between 3.58 (“the SMARTtest app was fun and entertaining to use”) and 4.48 (“I trusted the information (e.g. HIV/STI facts, referrals, etc.) presented on the SMARTtest app”).  100% said that they would use the INSTI for self-testing; 89% said they would use the INSTI for partner testing. | NR | NR | NR |
| **Baraitser 2019** | NR | NR | 1502 orders were placed; 1466 kits were dispatched.  67% chose HIVST, 34% chose self-sampling. | Test results were obtained for 57.2% of the HIVSTs and 53.9% of the self-samples. | NN |
| **Bauermeister 2015** | Participants completed a satisfaction survey which operated a 7-point Likert scale (higher number = higher satisfaction).  Descriptive statistics not reported; assumed to be mean (SD) given that t-tests were reported. | **Intervention**  Overall satisfaction = 6.16 (1.08).  Frustrating usability = 2.09 (1.27).  Recommend to friend = 6.00 (1.21).  Easy to use = 6.29 (.96).  Provides accurate info = 6.35 (.88). **  Likelihood of continuing use = 5.77 (1.30).  **Control**  Overall satisfaction = 6.00 (.77).  Frustrating usability = 2.19 (1.44).  Recommend to friend = 5.74 (.99).  Easy to use = 6.24 (1.01).  Provides accurate info = 5.74 (1.15). **  Likelihood of continuing use = 5.79 (.93).  **p<.01 | 130 people recruited. | 104 (80%) completed the 30-day follow-up. | NN |
| **Biello 2021a** | Cross-sectional survey | SUS = 71 (SD = 11.8)  73% were very satisfied with the app.  91% would recommend the app to a friend. | 11 | 100% retention | Participants used the app an average of 8 times (SD = 5) for an average total duration of 4 hour and 39 minutes. All functions of the app were used to some degree, the least used was the FAQ section which was only used by 4 participants. |
| **Biello 2021b** | Cross-sectional survey | 93% of participants stated that it wasn’t difficult to order an HIVST via the app.  87% of participants stated that the app was extremely or very helpful when ordering HIVSTs.  80% of participants who used an HIVST during the study (n=20) reported that HIVSTs would be convenient in the future. | 80 | 71 (89%) completed at least one follow-up assessment. | NN |
| **Chan 2021** | Cross-sectional survey | 74.3% of participants said they would be likely to use free HIVST with online counseling in the next 6 months.  76.0% of participants said that the HIVST was easy and 82.3% said it was convenient.  79.1% said HIVSTs could reduce embarrassment; 50.0% said they could help avoid stigma; and 84.0% said that they could improve privacy.  64.0% of participants stated that the online real-time counseling was important or very important for supporting new HIVST users. | 350 | 337 participants accepted an HIVST kit, 169 completed the HIVST-online service, 155 of whom were followed-up at 6 months.  143/168 who refused the HIVST-online service were followed-up at the 6 month mark. | NN |
| **Chasco 2021** | Qualitatively via interviews. | Participants felt that the service was convenient and was able to circumvent some of the barriers typically associated with testing. | 77 people were offered home kits, 42 (54.5%) accepted them resulting in 207 monitoring episodes (79 of which were using home kits). | NR | HIV samples were completed at 100% of the clinic visits and 83.5% of the self-samples. Adequate sample for creatinine analysis was obtained in 91.9% of the clinic visits and 81% of the self-samples. |
| **Cohen 2017** | NR | NR | 1460 people’s data were included (681 of which was collected following implementation of the intervention). | NR | The intervention appeared to be successfully implemented and resulted in shorter mean waiting times.  41.51% of participants opted into the intervention. |
| **Daniels 2016** | Participants completed a survey that asked for their most preferred method of communicating results and their level of comfort taking a picture of completed tests and texting their results. | **Most preferred method of sharing results**  E-mail = 37.8%  Text = 21.6%  In person = 16.2%  Taking and sending a pic of used test = 13.5%  In writing = 5.4%  Mailing in used test = 2.7%  By phone = 2.7% | NA | NA | NA |
| **De Boni 2019** | NR | NR | 17.,786 unique visitors to the study website; 7,300 questionnaires initiated; 4,800 questionnaires were eligible; 3,885 packages requested; 2,526 packages delivered | 542 packages returned | Participants who received an HIVST after starting the online process = 65% (success indicator of ≥60%).  Individuals who collected their HIVST from the pharmacy within two weeks of ordering = 38.5% (success indicator of ≥50%).  HIVSTs distributed during the first 12 months = 2526 (success indicator of ≥1000).  Proportion of HIVST results returned = 21.4% (success indicator ≥20%).  Proportion of positive tests followed-up with confirmatory testing = 88.2% (success indicator ≥50%). |
| **Elliot 2016** | NR | NR | 17,361 participants completed the baseline assessment.  7,872 (45%) were identified as being at risk of acquiring HIV.  11,127 (64.09%) clicked through for more information.  10,323 (93%) requested an HIVST kit. | 55% of those who requested an HIVST kit returned it. | 58% of the participants who were identified as being at high risk of acquiring HIV sought further information.  Blood tests were preferred but saliva tests were more likely to be returned.  82 people were newly diagnosed with HIV (1.4% of the participants who returned samples). |
| **Finkenflügel 2019** | NR | NR | 374 | 92.5% completed the 12-month follow-up. | The percentage of participants who reported data 27 to 30 days per study month decreased over time (p<0.001).  By the 12-month point, 37.7% of participants had at least one period of at least 30 days where they did not use the app.  PrEP adherence measured by app data and follow-up questionnaire were comparable for those on a daily regimen.  The median number of pills taken according to the app tended to be lower than reported in the questionnaire for those on an event-based regimen – this was significant at month 9 and the questionnaire was administered every three months. |
| **Fuchs 2018** | Qualitatively via focus groups. | Participants reported that the intervention provided additional support and security.  Participants reported that the intervention was unnecessary for those who were already fully adherent.  Staff found the intervention easy to implement. | 56 people enrolled in the study. | 52 (92.9%) completed the study:  3 people were unable to activate their email accounts.  1 person withdrew due to a technical error. | 18 participants (32.1%) opted for email over SMS.  80% of participants preferred weekly message frequency at the start of the week.  Participants who opted for email were less likely to reply (14% did not reply at all compared to 9% who opted for SMS).  4% of messages were not delivered due to an early rectified technical issue.  Email respondents took longer to reply. |
| **Gilbert 2017** | NR | NR | 868 people created accounts. | 15-25% discontinued at each stage.  318 (36.6%) participants submitted specimens.  96 (30.2%) retested within the study period. | 3.1% of participants had a positive STI diagnosis. 1.7% opted out of urine test.  5.8% opted out of HIV test. 5.0% opted out of syphilis test. 8.8% opted out of HCV test.  All participants who were received a positive test result received their results over the phone within 6 days. 90% confirmed they had received treatment.  76% of those with negative results were known to have clicked on their notification email but it is the others may have logged directly on to the website. |
| **Gilbert 2018** | NR | NR | 100 were contacted to take part | 73 participated | NN |
| **Gilbert 2019a** | NA | NA | NA | NA | NA |
| **Gilbert 2019b** | NR | NR | 19,497 | NA | For GCO, there were 1951 testing episodes across 1093 clients.  For the STI clinics, there were 39,357 testing episodes across 18,404 clients.  STI clinic clients had repeat tested 1.53 times/person year and GCO clients had repeat tested 1.87 times/person year (this was statistically significant).  Most GCO clients only tested through GCO. Of the 272 who tested at both GCO and an STI clinic, 87% were tested at a clinic first (44 of whom went on to test at a clinic again), 13% were tested through GCO first. |
| **Guinness 2018** | NA | NA | 126 patients were identified as being eligible for the study. 77% of patients were sent an email. | 78% of those send an email opened the email. | 12.4% of those send an email were linked to PrEP care, 91.7% of whom had a prescription filled. |
| **He 2018** | NA | NA | 500 HIV self-sample kits were distributed. | 430 (86%) of kits were completed and returned. | 16.3% of the returned kits tested positive for HIV. |
| **He 2019** | NR | NR | 957 tests distributed. | 378 tests were returned (2 were invalid).  255 participants accessed their results. | 65.9% of participants whose test was negative accessed their results. 100% of participants whose test was reactive accessed their results (n=7).  88.9% of the kits were dispensed between 9pm and midnight.  72.2% of the kits were dispensed in dorms, 27.8% were dispensed in teaching buildings. |
| **Hoth 2019** | NR | NR | 186 referrals resulting in 127 initial video calls. | 83 started PrEP and had enough time to measure follow-up within the study period. 60% of these were retained at 180 days. | There were 186 referrals which resulted in 127 initial video-consultations (all completed within 40 days of the referral; 78% of these were completed within two weeks of referral; and 53% within one week).  Retention was 60% (50/83) at 180 days (for clients who had enough time for follow-up to be measured).  Completion of blood tests was 96%.  1 client was newly diagnosed with HIV at baseline.  Of the 167 eligible creatinine visits, 98% (n=164) were complete. |
| **Hottes 2012** | Qualitatively via interviews. | **Perceived benefits of internet based testing:**  Anonymity, immediate access, 24hr availability of internet and extended/flexible hours of lab sites, and standardised service, controlled by client.  **Concerns re: internet based testing:**  Reluctance to provide personal info online; distrust of security of data provided online; ensuring comprehensive pre-test counselling; support for those waiting for test results and receiving positive results.  Most participants would use the internet service in the future or recommend it to others. | NA | NA | NA |
| **Huang 2016** | Cross-sectional survey | **Ease of using self-test**  Very easy (58%)  Easy (39%)  Neutral (4%)  Hard (0%)  Very hard (0%)  **Testing preference**  Prefer self-test kit (44%)  Somewhat prefer self-test kit (25%)  Neutral (12%)  Somewhat prefer clinic (16%)  Prefer clinic (4%) | 62,820 people potentially saw the promotion.  There were 11,939 unique visitors to the study website (an average of 284 per day).  Click-through rate of 2.8%.  238 people were interested in participating in the study. 122 were eligible. | 81 (66.4%) of participants confirmed that they had received an HIVST kit and completed the follow-up. | NN |
| **Hughes 2021** | Qualitatively via interviews | Most participants were extremely satisfied with the service and reported having to “overcome scepticism” as the service appeared too good to be true or a departure from the care they were used to receiving.  Participants found the web-based nature of the service convenient and felt it was able to strike a balance between efficiency (simplicity, speed, convenience) and “humanity” (personalised, responsive, feeling of a connection/care). | NR | NR | NR |
| **Jackman 2018** | Cross-sectional survey | Effect of ePHR on communication of STIs: 63.6% very helpful/helpful.  Effect of ePHR on confidence in partner's results: 66.4% very helpful/helpful.  ePHR will make it easier for talks re: STI testing: 55.6% strongly agree/agree.  ePHR make it easier to check-in on partners: 55.1% strongly agree/agree.  Soliciting a partner's ePHR will be awkward: 24% strongly agree/agree.  Confidence in sharing positive STI through ePHR: 16.7% very easy/easy.  Belief partner will initiate STI talk earlier: 52% strongly agree/agree.  Only use ePHR when distrusting of partner: 26.3% strongly agree/agree.  Suspicious of partner who is unwilling to share: 75.7% strongly agree/agree. | NA | NA | NA |
| **Jin 2018** | NR | NR | 1,015 people applied for an HIVST kit; 879 (86.6%) were eligible. | 77.7% of participants who received a test submitted a photo of their results. | 40% of participants had never been tested for HIV before.  14.3% of the people who submitted their results were found to be living with HIV.  72.4% of the people newly diagnosed with HIV were receiving treatment within 1 month of their test.  42.9% of those who uploaded a photo of their test were first time testers and among the first time testers, 18.8% were diagnosed with HIV. |
| **Kecojevic 2018** | NA | NA | NA | NA | NA |
| **Knight 2019** | Qualitatively via interviews | Participants preferred GCO because it was convenient, private and they felt that they had more control.  Participants noted that it improved access for rural patients and that they preferred getting results online rather via email or over the phone.  Almost all participants said that they would use the service again in the future. | NA | NA | NA |
| **Koester 2020** | Qualitatively via interviews | Participants found presenting to a clinic to testing or to drop of specimens acceptable. The authors note that Nurx has a variety of sites with long opening hours. | NR | NR | NR |
| **Lee 2020** | NA | NA | NA | NA | NA |
| **Lessard 2019** | Qualitatively via focus groups. | Overall acceptability appears good.  Potential benefits of the service are that it could allow better patient notification, customized linkage to care, and transferrable data.  Potential risks could be privacy and confidentiality, changes in sexual behaviour, and fairness. | NR | NR | NR |
| **Liu 2021** | Cross-sectional survey and focus groups. | Median SUS score was 85/100 at week 4 and 80/100 at week 8. 84% reported that they were likely to use the app again in the future. | 20 | 1 lost to follow-up | Among 1198 expected doses, 80% were confirmed, 6% were self-reported, and 14% were missed.  Median PrEP adherence was 91%. |
| **MacGowan 2019** | NR | NR | 2665 | Retention was >54% at each follow-up. | A significantly higher proportion of participants in the HIVST group reported testing 3 or more times during the trial than in the control group (76.6% vs 22.0%; p<0.01).  The largest difference in HIV diagnoses was during the first three months (0.9% HIVST vs 0.1% control; p<0.01). |
| **Maksut 2016** | Cross-sectional survey | All participants reported that they would like to participate in at-home HIV testing with peer counseling via video chat in the future and that they would recommend this to a friend. 72% said they would prefer this mode for their next HIV test. | 23 people agreed to participate, 20 completed the HIV testing appointment. | 18 (90%) participants were retained for follow-up. | NN |
| **Manavi 2017** | NA | NA | 5,130 kits were requested. | 3,099 (58.4%) kits were returned. | Kits delivered to homes were more likely to be returned (60.6%) than clinics (56.4%) and pharmacies (44%). |
| **Menza 2021** | Follow-up email | Participants said that the service was convenient and enhanced privacy. | 233 participants ordered a total of 248 kits. | NA | 22% of the participants lived in rural zip codes.  73% of the 150 kits assigned to the study to begin with were ordered within the first 24 hours of implementation. |
| **Mitchell 2018** | Cross-sectional survey.  SUS and 4-point Likert scale (1= not at all; 4 = extremely).  Mean (SD) reported. | **SUS = 68.25 (15.10)**  Overall satisfaction = 2.80 (.63).  Usability on a daily basis = 3.50 (.53).  Recommend to others = 2.70 (.82).  User-friendliness = 2.80 (.79).  Difficulty learning to use = 1.20 (.42). | 10 people participated in the study. | 100% retention. | Daily doses were registered 91% of the time. Among these, 88% involved the camera.  40% of participants didn't miss any days; 40% missed 1-5 days; 10% didn't log a dose on six days and 10% for 12 days.  70% of the participants responded to all of the daily surveys. |
| **Page 2019** | NA | NA | 550 kits (275 each of MT and DBS) were requested online. | 98.8% of the DBS were returned and 55.7% of the MT were returned. | 96 MT were not processed, 62 of which were due to insufficient sample.  21 returned DBS were not processed, 2 of which were due to insufficient sample.  5.4% false positive rate for MT compared to 0% for DBS. |
| **Phanuphack 2018** | NA | NA | 564 participants were recruited:  202 selected the offline arm.  158 selected the hybrid arm.  211 selected the online arm. | 100% of the offline group completed testing.  94.3% of the hybrid group completed testing.  92.4% of the online group completed testing. | Percentage of each arm that were first time testers: 42.4% in offline; 18.1% in hybrid; 47.3% in online.  13% of the people in the offline arm were diagnosed with HIV, 3.4% in the hybrid arm, and 15.9% in the online arm. |
| **Polilli 2016** | NA | NA | The authors used approximations:  6000 visited the website; 5000 completed the risk calculator; 3500 booked a test; 3,046 presented for testing. | Retention was covered in the uptake column. | NN |
| **Refugio 2019** | Cross-sectional survey | >85% agreed that PrEPTECH was a better way for GBMSM to access PrEP (measured at 90- and 180-day follow-up).  88% of participants reported that PrEPTECH was very or extremely easy to use.  >85% of participants said that they trusted PrEPTECH. | 401 people created an account.  44 completed the consent process. | 25 participants completed baseline; 21 completed follow-up; 16 were interested in continuing PrEP after the study ended; 11 were confirmed to have accessed PrEP after the study ended. | NN |
| **Rosengren 2016** | Cross-sectional survey | 93% of participants found the HIVST kits easy or very easy to use.  77% of participants preferred or somewhat preferred self-testing over in person testing. | 4,389 visitors to the website, 333 requested a test. | 125 participants completed the online survey.  56 (45%) completed the follow-up survey. | 4% of the people who completed the follow-up survey tested positive for HIV; all of whom were linked to care. |
| **Salway 2019** | NR | NR | 352 | NR | NN |
| **Siegler 2019** | Cross-sectional survey | SUS mean (SD) = 76.91 (18.4).    85% indicated that they would prefer to use the service in place of a standard visit.    40% reported that they would be more likely to stay on PrEP if the service was made widely available.  **Acceptability of features**  Packaging and mailing results (89.1% said acceptable or very acceptable).  Urine specimen collection (92.7% said acceptable or very acceptable).  Rectal swab specimen collection (87.3% said acceptable or very acceptable).  Finger prick and blood collection in micro-tube (58.2% said acceptable or very acceptable).  Finger prick and dried blood spots (69.1% said acceptable or very acceptable).  It is worth noting that, for the latter two, the unacceptable/very unacceptable response was <20% for each. | 58 participants. | 3 participants were lost to follow-up. | 1 participant had an insufficient volume of blood for remote testing.  1 participant’s rectal test sample not done correctly so could not be analysed.  Of the 57 participants whose data was available, 4 required and received standard face-to-face care instead due to 2 being unable to prick their finger and 2 having insufficient specimen collections.  93% were able to have their prescription renewed based on the online service. |
| **Stekler 2018** | NR | NR | 48 | 40% of the participants in the intervention group attended the 3-month follow-up compared to 87% of the control group. | NN |
| **Stephenson 2020** | Cross-sectional survey | Satisfaction was high (overall satisfaction = 98.3%):  98.3% thought the counsellor was friendly; 100% thought the counsellor was knowledgeable; 98.3% thought the counsellor was experienced; 98.3% thought the counsellor was professional; 78.3% found the intervention easy to use; 73.3% thought the intervention was good quality; 98.3% thought the HIVST was easy to use; 93.3% thought the HIVST was easy to interpret; 89.8% would be willing to repeat the session; 81.3% would be willing to recommend the session; 98.3% would be willing to recommend home test. | 202 | 100% of the control group ordered HIVST, 91% reported their HIV test results via the study portal. 2 participants reported a preliminary positive result and were contacted within 48hrs and linked to care within 30 days.  48% of those randomised into the intervention group opted to receive the intervention - all of whom ordered HIVST and received video-chat counseling. The HIVST was performed in the session in 96% of cases. All had a non-reactive result. | NN |
| **Sullivan 2017** | Cross-sectional survey | 88% found level of detail useful/very useful.  81% found assessment recommendations useful/very useful.  66% felt the app content helped them to stick to HIV prevention plan.  71% found the app to be a good balance of personal and professional language.  90% found the information easy to understand.  86% found the app to be secure.  85% found the password/PIN offered sufficient protection.  84% found the app name and icon not to be readily associated with HIV prevention.  Composite usability score was 73.4 (above average) | 919 survey responses, 309 of which were eligible. Final enrolment was 121. | 81% of participants completed the 4-month evaluation. | NN |
| **Syred 2019** | NR | NR | 6,253 users ordered 7,550 tests prior to implementation; 7,772 users ordered 9,785 tests following implementation | NA | When compared to data for the same time period prior to implementing the ‘choose to test’ service, the positivity rate for gonorrhoea and chlamydia tests was unchanged. Too few HIV and syphilis tests were completed to analyse.  GBMSM were offered genital, rectal and oral chlamydia and gonorrhoea tests plus HIV and syphilis testing - in 17.2% of orders, people removed tests.  BME users were offered chlamydia, gonorrhoea and HIV testing, in 77.9% of order, people added tests. |
| **Wang 2018** | Cross-sectional survey | Satisfaction with logistics of implementation = 89.5%.  Satisfaction with performance of HIV testing admin = 96.5%.  Satisfaction with usefulness of the HIVST-OIC in helping understand HIV testing (86.7%).  Satisfied with usefulness of HIVST-OIC in preparing them to take up such a test (80.3%).  Perceived effectiveness of intervention in reducing risk behaviours (71.4%).  Use the service again (78.5%).  Recommend to a friend (75%).  Would pay for HIVST-OIC in the next six months (47.8%). | 430 | Loss to follow-up:  10.7% intervention; 6% control | NN |
| **Whiteley 2020** | NA | NA | NA | NA | NA |
| **Wilson 2017** | Cross-sectional survey | 71% of participants found the service acceptable. | 2,072 were randomised | 1,739 (83.9%) completed follow-up. | NN |
| **Witzel 2019** | Cross-sectional survey | 98% found the instructions easy, 97% found the HIVST simple to use, 97% had an overall good experience. | 1370 people registered; 1035 (76%) enrolled | Of the 631 randomised to receive an HIVST at baseline, 78% completed one of the follow-up surveys. | 97% of the participants in the baseline HIVST arm received an HIVST and 90% had used it. |
| **Witzel 2021** | Cross-sectional survey | 97% found the instructions easy, 97% found the HIVST simple to use, 100% had an overall good experience. | 118 | Randomisation 1:  Baseline test = 72% retention rate  No baseline test = 41% retention rate  Randomisation 2 (those with baseline test were randomised to receive a repeat test or not):  Repeat test retention rate = 100%  No repeat retention rate = 88% | In trans men, HIV testing uptake was significantly higher in the baseline test group (95%) than in the no baseline test group (29%) (p<0.001). Trans people in the repeat test group had a 3 times higher rate of repeat HIV testing compared to no repeat test group (p<0.0001). |
| **Woywodt 2014** | Cross-sectional survey | 84.8% of participants rated the service as good or v good. | 295 returned questionnaires (response rate of 45%). | NA | NN |
| **Wray 2018** | NR | NR | 65 | 12.3% of participants withdrew before month 7 | Monthly survey completion rate = 89%.  93.1% of the sample used the study-provided HIVST. |
| **Xia 2018** | NR | NR | 3,092 packs were distributed. | 1,977 (63.9%) samples were mailed to the lab, 1,911 (96.7%) were eligible for analysis.  65.4% of the people who submitted a sample accessed their results online, this was higher for the people whose HIV test was reactive (83%). | 7.1% of the samples tested positive for HIV. |
